# Informing Epidemic Control Strategies: A Spatial Metapopulation Model Incorporating Recurrent Mobility, Clustering, and Group-Structured Interactions

**DOI:** 10.64898/2026.04.08.26350398

**Authors:** Michael Lazarus Smah, Anna Seale, Kat Rock

## Abstract

Infectious disease dynamics are strongly shaped by human mobility, social structure, and heterogeneous contact patterns, yet many epidemic models do not jointly capture these features. This study develops a spatial metapopulation epidemic model incorporating recurrent group-switch interactions to represent real-world transmission processes. Building on the Movement–Interaction–Return framework, the model integrates household structure, age-stratified contacts, and mobility between locations within a single SEIR framework. Using UK demographic, mobility, and social contact data, the model quantifies how within- and between-group interactions, mobility rates, and location connectivity influence epidemic spread. Both deterministic and stochastic simulations are implemented to analyse outbreak dynamics, variability, and fade-out probabilities for COVID-19–like and Ebola-like infections. Results shows that highly connected locations drive faster transmission, earlier epidemic peaks, and greater difficulty in containment, whereas larger but less connected locations tend to produce slower, more localised outbreaks despite their population size. Comparative analysis reveals that COVID-19–like infections spread rapidly and remain difficult to control even under interventions, while Ebola-like infections exhibit slower dynamics and are more effectively contained, particularly under targeted measures. Non-pharmaceutical interventions, particularly widespread closures, substantially reduce infections, hospitalisations, and deaths, although effectiveness depends on timing and pathogen characteristics. These findings highlight the importance of integrating mobility, clustering, and demographic heterogeneity to inform targeted and effective epidemic control strategies.

## 1 Background

Infectious disease outbreaks can lead to epidemics and pandemics, which directly impact health and, more broadly, can have substantial economic and social impacts. Although the exact pathogen and starting location of outbreaks of pandemic-potential are unknown, the World Health Organization has listed priority diseases through which a future pandemic could occur, including COVID-19, Crimean-Congo hemorrhagic fever, Ebola virus disease and Marburg virus disease, Lassa fever, Middle East respiratory syndrome (MERS) and severe acute respiratory syndrome (SARS), Nipah and henipaviral diseases, Rift Valley fever, Zika and Disease X [1]. Some of these priority diseases do not possess all the characteristics expected to cause a pandemic, but are considered to have that potential within a (short) period of evolution.

There have been outbreaks of some priority diseases; some were contained locally [2, 3], others led to epidemics [4]. Most recently, COVID-19 led to a pandemic with its first reported case in Wuhan, China, in December 2019 [5]; in response, various countries implemented NPIs. Some intervention strategies were helpful; however, their effectiveness has faced scrutiny [6]. The connection of China to other parts of the world probably contributed to the rapid spread of COVID-19 [7]. In this context, it is important to explore factors that increase the risk of epidemics and pandemics, rather than simply the biological characteristics of these pathogens with pandemic potential.

In the most recent pandemic of COVID-19, the infection spread quickly around the world. The first incidence of COVID-19 was recorded in China in December 2019. By February 21, 2020, only three months later, 28 countries on different continents were already affected [5]. The 2009 H1N1 influenza (swine flu) pandemic followed a similar pattern. With initial cases in Mexico and the United States in April 2009, H1N1 then spread around the world within weeks [8]. Both pandemics showed how quickly respiratory diseases spread in the connected world.

Many factors influence the spread of outbreaks, unrelated to the pathogen. These include people’s usual mobility, their connectedness in and outside of the household, as well as the nature of interactions. The interplay of these factors and pathogen characteristics, including mode of transmission, transmissibility and clinical symptoms, will also influence transmission. The impact of interactions within households and subsequent dispersion to different locations during daytime that could result in transmission is an interesting area that could help provide a framework to quantify priority groups of people, and their locations before and during outbreaks, for adequate planning and intervention.

Considerable efforts have previously been made to model the dynamics of infection within households. Some of these models [9–11] provide a framework that allows households to be categorised according to their epidemiological states (susceptible, infected, recovered, etc) and have provided a useful way of accounting for disease transmission dynamics within households.

A mathematical metapopulation model in which a population is partitioned into subpopulations that each undergo an epidemic process has been used (e.g., [12–16]) to capture the effect of mobility on epidemic spread from one location to another. The metapopulation modelling approach often utilises ordinary differential equations (ODEs) for mathematical tractability; however, equation-based models (EBMs) like the ODEs usually make simplifying assumptions regarding mobility patterns and contact mixing processes. These simplifying assumptions, however, don’t fully capture the complexity of human mobility and other complex characteristics, which may result in less robust predictions. Many individual-based models (IBMs) have been used to incorporate complex human characteristics in epidemic models [17–20]; however, the computational nature and the large number of parameters required for IBMs and the complexity of fitting to data, among other factors, makes it challenging to give analytical insights into the effect of the parameters[21, 22]. Despite the seeming simplification of some of the classical ODE modelling approaches and the challenges in the use of the IBMs to give analytical insights about the parameters governing epidemic spread, their importance and usefulness in understanding and thus informing interventions to mitigate infectious disease spread cannot be overemphasised.

In order to retain the property of capturing complex human interaction while allowing for an analytical understanding of epidemic processes, a theoretical framework based on probabilistic discrete-time Markov chains [23] has been explored to derive equations that describe how a susceptible individual becomes infected when in contact with an infectious individual. To improve the realism of recurrent human mobility where individuals make daily mobility to work and back home after work to epidemic spread, the Movement-Interaction-Return (MIR) epidemic model [24] was proposed following the probabilistic discrete-time Markov chains [23] to account for the fundamental mixing processes driving epidemics, as well as the impact of spatial distribution of populations and commuting mobility patterns where individuals leave their homes at the beginning of each day to their places of activities and return after the work-day interaction.

Studies on the MIR models [21, 22, 24–30] provide a formalism that makes it possible to analyse the epidemic risk in systems that display recurrent mobility patterns in a reliable, analytically and computationally efficient manner. In the MIR framework, it is found that there is a regime where mobility hinders the spread of pathogens, which appears counterintuitive. These results have been supported by the findings in the literature [31]. The MIR models typically describe the dynamics of infection spread in terms of the probability that each node (which could be a location or an individual, depending on the study) progresses through different infection states.

A PubMed search for *“age-structured metapopulation epidemic model”* (May 1, 2005 – May 1, 2025) returned four original modelling studies that explicitly developed and applied age-structured, spatially-distributed epidemic models, providing data-driven analyses of how age structure and spatial coupling shape transmission and the effectiveness of interventions.

Coletti et al. [32] developed a stochastic, data-driven metapopulation model for Belgium, integrating age-specific contacts and mobility to assess lockdown and phased exit strategies. Their model revealed that cutting back on leisure and other non-essential interactions notably impacts resurgence risks more than reopening schools, highlighting how contact settings, age demographics, and mobility influence epidemic rebounds. Chinazzi et al. [33] developed a multi-scale modelling framework that is structured by age, accounts for multiple strains, and explicitly incorporates vaccination, mobility, and NPIs. Their findings indicate that spatial heterogeneity and the timing of control measures play crucial roles in determining the establishment and regional prevalence of variants. Su et al. [34] employed an age-structured metapopulation model across different Chinese provinces to assess rubella burden and evaluate immunisation strategies. They used serological and surveillance data to demonstrate how demographic make-up and inter-provincial connectivity influence disease burden and the projected outcomes of interventions. Metcalf et al. [35] utilised provincial data from Peru in a metapopulation framework to illustrate that weak spatial coupling and extended fade-outs can shift infection age distribution upwards. This occurs because prolonged fade-outs prevent the depletion of susceptible individuals through infection and recovery, resulting in older individuals remaining susceptible, which increases the risk of congenital rubella syndrome in less connected regions—an effect driven by the interaction between spatial structure and age-specific susceptibility.

Although these original studies integrate age structure and spatial (metapopulation) coupling—and many leverage high-quality empirical data for calibration—none of the four explicitly model households as discrete mixing units with within-household transmission linked to individual mobility and cross-group contacts. A search for *“age-structured metapopulation epidemic model with households”* returned no results, highlighting the scarcity of models that jointly capture household-level transmission, demographic structure, and spatial mobility. This omission is consequential: households are a principal micro-scale setting for infection transmission and for bridging demographic groups (for example, school children, working adults, and older adults often cohabit), and household exposures can seed infections that are then transported across locations by the heterogeneous mobility of different household members. Models that treat age structure and mobility without representing household composition may therefore understate short-term amplification, mischaracterise cross-group seeding events, and misestimate the downstream spatial spread and intervention impacts.

To improve predictive realism and intervention design, modelling work should aim to unify age-stratified contact matrices, household structure (e.g., household size distributions), and metapopulation mobility (both routine commuting and long-range travel) within a single modelling framework. Such multi-scale models would allow direct quantification of (i) how intra-household transmission couples otherwise distinct demographic and mobility strata, (ii) the contribution of household composition heterogeneity to local outbreak persistence and regional seeding, and (iii) the differential effectiveness of interventions (e.g., household isolation, targeted vaccination, mobility restrictions) that act at household, community, or regional scales. The scarcity of original studies that simultaneously incorporate these elements highlights a clear and important gap in the infectious-disease modelling literature.

It is important to develop models to consider the recurrent group-switch behaviour which we define as a phenomenon accounting for households made up of different groups (age groups, work groups, and other risk groups) disintegrating during work time with each household member going to different locations and interacting with different groups before returning home after work day thereby switching between the household group and work-groups every day. The specific household interaction, the probability of within/between group interaction, the average number of total daily contacts captured by social contact data, and the possibility of groups forming clusters of interaction within each location are important constituents that could result in variations observed in the spread of diseases across different population groups and locations. Incorporating all of these in a single model will help to inform control strategies by understanding how a particular pathogen may spread in different settings, depending on the heterogeneous characteristics of the people and locations.

The concept of *semi-random mixing*[36] have been introduced, which allows the incorporation of the number of clusters characterised by the formation of smaller groups from which people randomly choose their daily contacts for interactions. The concept allowed the incorporation of multi-level interaction for both household and work-time interactions in literature [37], particularly, it disentangled the probability of mobility, the average number of daily contacts, and the duration of contacts within and outside the households from the biological infectiousness and recovery rates of pathogens, giving us insights into the impacts of each parameter in the epidemic model and offering flexibility to the counter-factual analysis of intervention strategies.

The multi-level interaction model was extended in [31] to account for social heterogeneity in epidemic modelling. The model captured the switching between household groups and work-time groups; however, it did not account for the spatial distribution of people. In the framework, key quantities were defined: the source-to-sink, the source, and the sink reproduction numbers, and showed that these quantities are informative as part of modelling pathogen spread in heterogeneous populations. These quantities enabled me to quantify the contribution of different groups in transmission and reveal the risk of groups in relation to other groups.

This study will further the understanding of the contexts that increase epidemic and pandemic risk, as well as describe the relative potential effectiveness of different intervention strategies. This will be done by developing a spatial metapopulation modelling approach that incorporates the concepts introduced in the literature [31, 36, 37]. The approach factors in the manner in which individuals within the same household disperse to various locations and interact with various groups of people before returning home.

### 1.0.1 Research Questions

The following research questions guide the study:

a. How does introducing infections in different locations affect the final size and timing of the outbreak?
b. What is the level of uncertainty in outbreak outcomes due to the stochastic nature of the model?
c. How likely is it that a large outbreak could have been avoided without non-pharmaceutical interventions (NPIs), i.e., what is the risk of disease fade-out versus major outbreaks?
d. How do NPIs (e.g., travel restrictions, school/work closures) impact outbreak size and dynamics?
e. How would results differ for a moderately low-severity but fast spreading, COVID-like pathogen where many infections go undetected compared to a high-severity but low spread, Ebola-like pathogen?

## 2 Methods

### 2.1 Deterministic modelling approach

The metapopulation model in this work is in the form of the Movement-Interaction-Return (MIR) epidemic model [24] represented by the network diagram in Figure 1. The model is developed for an arbitrary group *g* in location *l*. This initial model will be applied to any group in the population.

**Figure 1.**
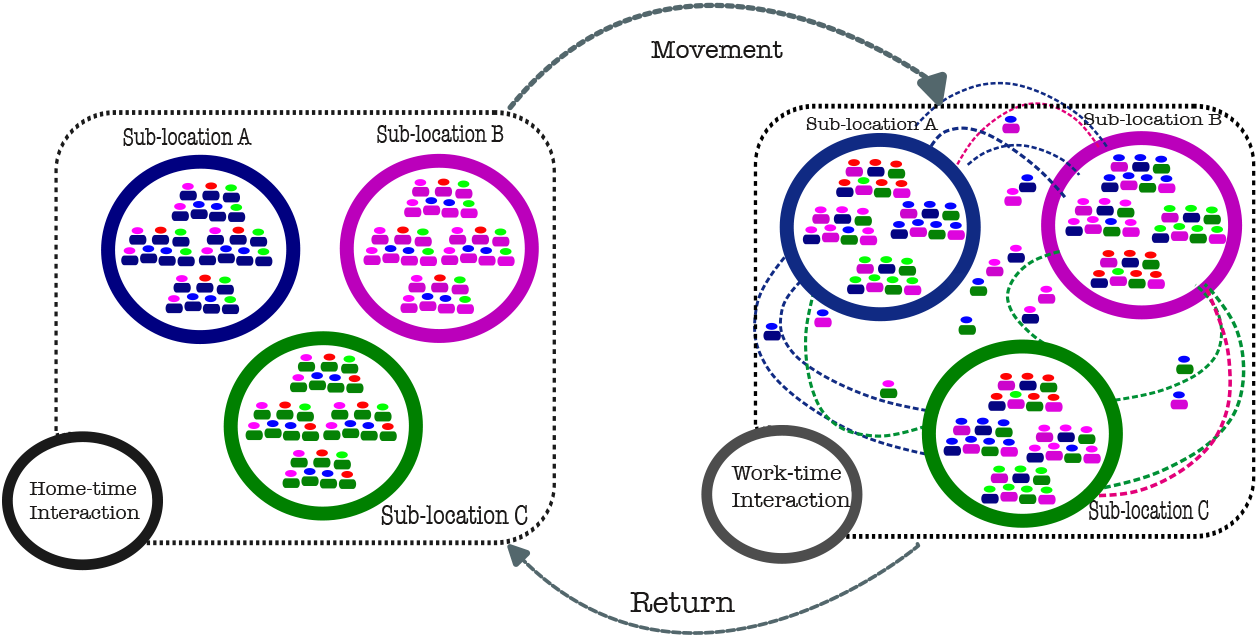
Network diagram for a metapopulation of three locations labelled (*A, B* and *C*). The colours of the head indicate socio-demographic structure, while the colours of the bodies signify location. Clusters during home-time interactions are dominated by body colours, regardless of head colours, indicating the usual interactions within households, while clustering during work-time interactions is dominated by the head colours, regardless of the body colours, signifying the coming together of individuals from different locations during working hours.

Let the number of individuals belonging to group *g* who are residents of location *l* be denoted by 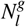. Let the proportion of individuals in this group that would move out of their households to work/school be 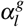. Assuming that, regardless of the location of the interaction, the average household neighbours of an individual in group *g* who are members of group *ġ* is 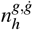, average external connections per individual 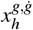. Also, let the average cluster neighbours at work of an individual in group *g* who are members of group *ġ* be 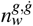, and the average external connections per individual at work is 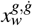. Let the average household contacts of an individual in group *g* who are members of group *ġ* be 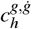, and the work/school contacts be 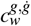.

If the number of individuals belonging to group *g* who are residence of location *l* that work/school at location *m* is by 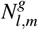, then the probability that any person in group *g* living in any location *l* will choose another location *m* as their destination when they make mobility decision is given following MIR literature [21, 22, 24–30] as:

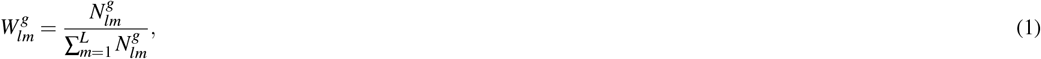

where *L* is the number of locations in the model. Here, *l* = *m* corresponds to the probability of moving to work/school in one’s location of residence.

At each time step *t*, an individual of group *g* will move from their households in location *l* to location *m* with a probability 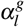 or remain within their households with probability 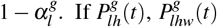, and 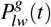 are the forces of infection for home-time interaction at home, work-time interaction at home, and work-time interactions at work/school respectively, the total force of infection for group *g* who are residents of location *l* is given, following similar expression for a single location in the literature [31] as:

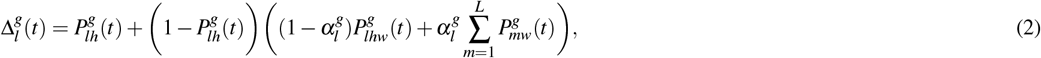

where:

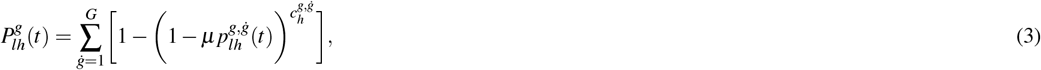

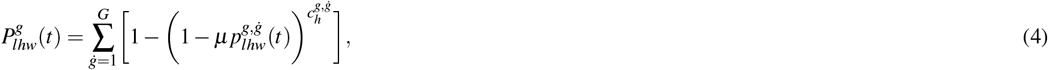

and

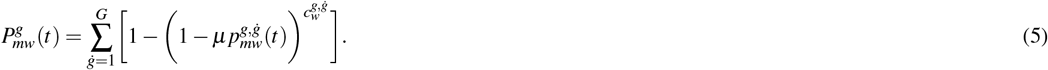

The probability that a person in any group *g* will meet an infected individual in group *ġ* during home-time and work-time interactions is given respectively as:

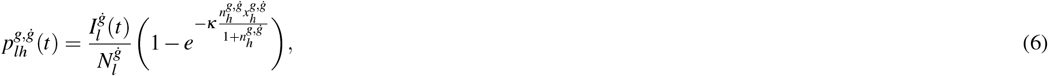

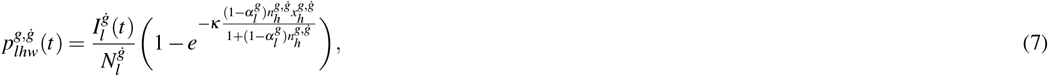

and

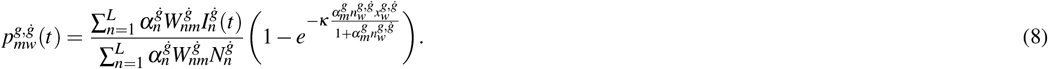

For *κ* being a coupling constant defined in the literature [36]. Linearised forms of equations (2) - (5) are given as:

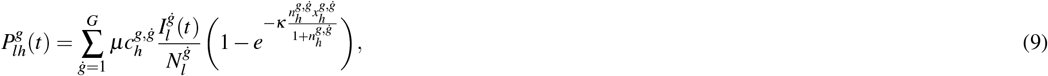

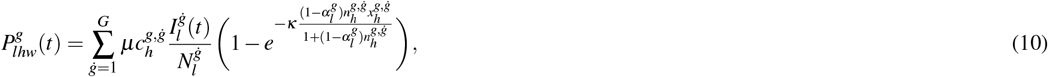

and

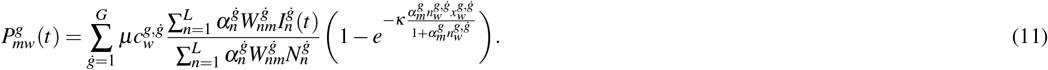

As epidemic processes continue in the population, accompanied by human activities, transmission to individuals in different groups and locations happens.

The mixing pattern in this framework that captures multi-level heterogeneity for both groups and locations makes it possible to account for specific within and between groups, and the within and between locations transmission.

The total force of infection in Equation 2 accounts for the transmission risk to members of the group *g* whose residence is in location *l* of the members of the group (when *g* = *ġ*) or other groups (when *g* ≠ *ġ*), including the transmission that occurs in their residence location (when *l* = *m*) and other locations (when *l* ≠ *m*). The equations representing the dynamics of disease transmission for the “Susceptible-Exposed-Infected-Recovered” (SEIR) type infection represented in Figure 2, where members of the group *g* whose residence is in location *l*, who are not infected but are prone to infection referred to as susceptible 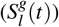 become infected with probability 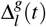 and remain exposed 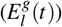 for a duration commonly termed as a latency period depending on the pathogen. An exposed person becomes infectious 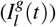 at the end of the latency period with a probability *σ* and can transmit infection to other susceptibles except they are removed/recovered 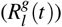 with a probability *γ* during which they cannot contribute to infection and cannot be reinfected.

**Figure 2.**
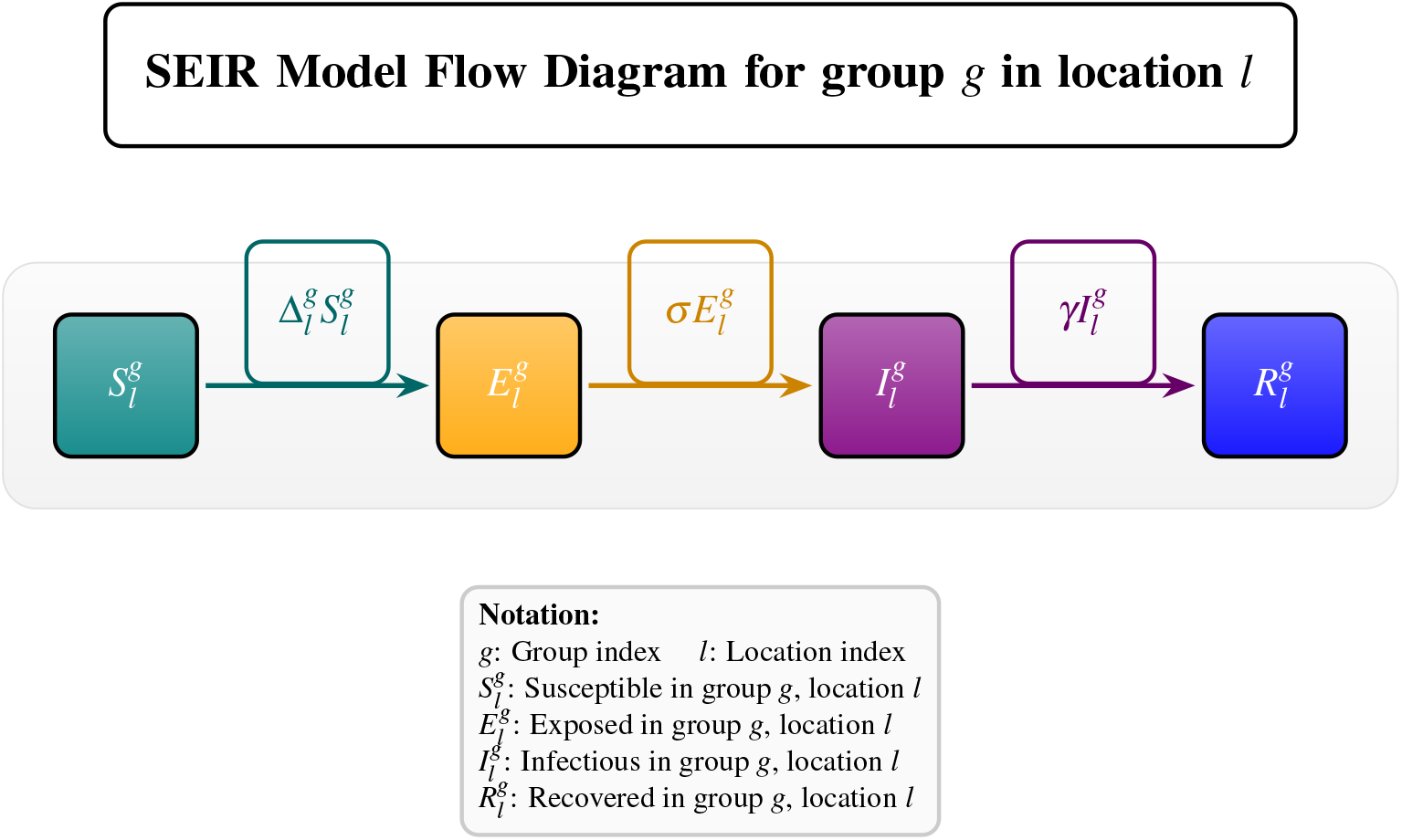
Schematic of the location-specific SEIR model for group *g* in location *l*. This shows the progression of infection from susceptible to recovery and the daily probabilities with which these transitions occur.

The discrete-time equations for the SEIR-type infection for any group *g* are then written as:

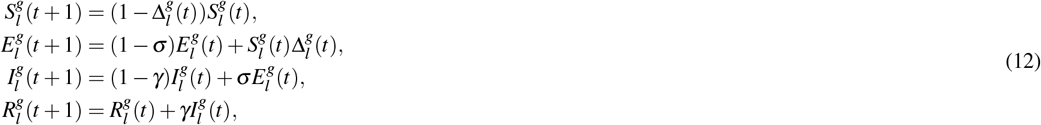

where 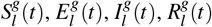, are the numbers of the susceptible, exposed, infectious, and recovered (removed) individuals at time *t* respectively, and 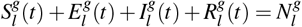.

### 2.2 The stochastic modelling approach

Stochastic models, unlike their deterministic counterparts, incorporate randomness and uncertainty into the modelling process. These models recognise that real-world systems often involve inherent variability and unpredictability in the input parameters and outcomes. Stochastic models use probability distributions to represent uncertain variables and generate a range of possible outcomes rather than a single deterministic prediction.

To implement the stochastic simulation of this framework, the same setup in equations (2)—(8) is adopted. The main difference is that the epidemic events occurring at each time step in equation (12) are implemented using binomial trials. For the SEIR-type dynamic, the events are infection, loss of latency, and recovery. The infection event in group *g* and location *l* at any time *t* is implemented using a binomial distribution with the number of people in that compartment and the daily probability of the event happening per person in that compartment. The daily SEIR model events are thus given by:

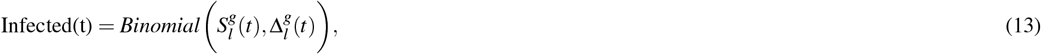

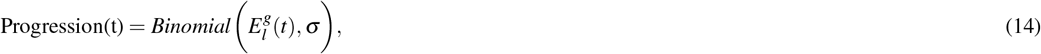

and

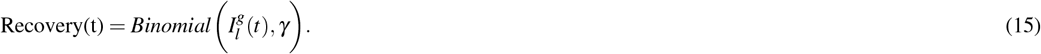

The discrete-time equations for the stochastic models are written as

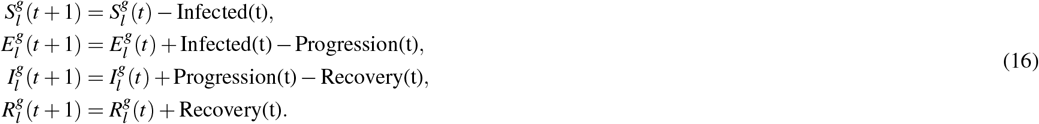

### 2.3 Data and parameter values

For numerical simulation, two pathogen-dependent parameter sets were used: the COVID-19-like infection parameters and the Ebola-like parameters. The social contacts and some of the COVID-19-like infection parameters were used as in the literature [31].

#### 2.3.1 Demographic mixing data

Publicly available UK social contact (POLYMOD) data [38] that shows the average number of contacts per day, within and between age groups, is used. The data have been extracted for age groups (6–24, 25–44, 45–54, 55–64, 65–74, and 75 plus). Two sets of the social contact data have been extracted: one for household mixing, representing *c*_*h*_ across age groups, and the other for non-household (work/school) mixing, representing *c*_*w*_ across age groups. The summary of the extracted contact network data is in Figure 3. The probability of leaving households to work/school is uniformly set for all age groups as *α* = 0.72.

**Figure 3.**
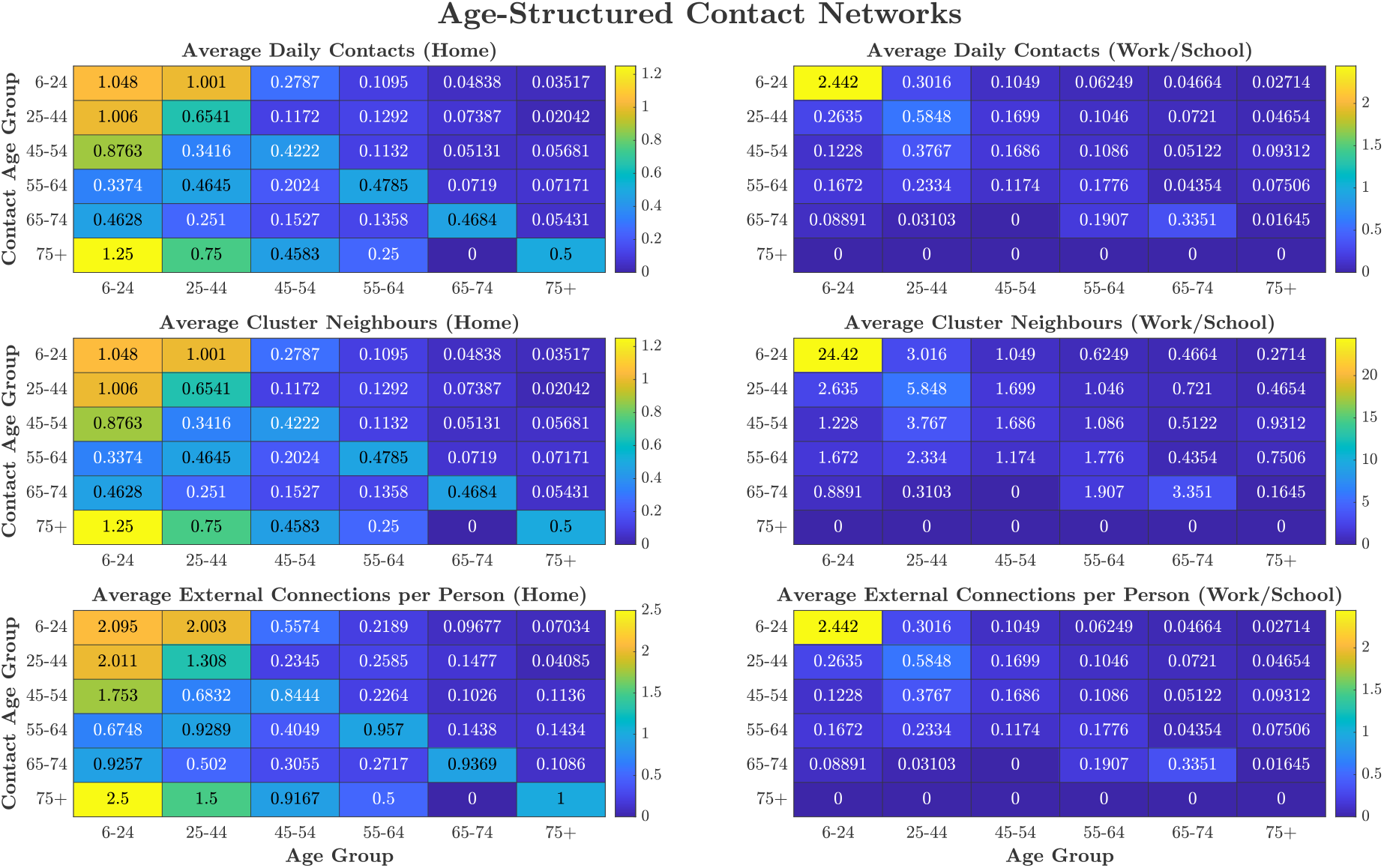
Age-structured contact networks derived from UK social mixing data illustrating average daily contacts, cluster neighbours, and external connections for home and work/school environments.

#### 2.3.2 Movement data

We sourced the publicly available UK census data for 2011 [39], which shows the number of individuals in each local area who have their work address in each of the local authority districts within the West Midlands. These information are used to construct a synthetic origin-destination (OD) matrix for six age groups and ten locations (Fig. 4), which are used to estimate the likelihood that members of any age group with residence in any location would normally choose any other location as their destination during work/school (computed using equation (1)). These locations are the places of residence and do not differentiate between neighbourhoods, schools, and workplaces. We assume that when people move at the start of each time step, their destinations are the usual work/school locations. The population distribution for 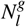 is computed from Figure 4. The total OD matrix of all individuals across all locations is given in Figure 5. The example of the network and the weight of interaction between locations is in Figures 6 (age group 75+) and 7 (all ages). For each location, we computed the percentage of the entire population who are originally from the location (Origin), the percentage of the entire population who normally go to work/school within their location of origin (Residents), the percentage of the entire population who leave their locations of origin to work/school in other locations (Out-flow), and the percentage of the entire population who visit each of the locations for work/school (In-flow). The summary of this information is in Table 1 and Figure 8. We used the network coupling constant as fitted in the literature [36] as *κ* = 0.6031. Except where indicated, the parameter values described above are used for simulation and referred to as the baseline values.

**Table 1.** Population Percentages by Location for Origin, Daily Out-flow, Daily In-flow, and Daily Residents.

| Location | Origin (%) | Out-flow (%) | In-flow (%) | Residents (%) |
| --- | --- | --- | --- | --- |
| 1 | 6.46 | 0.79 | 0.36 | 5.67 |
| 2 | 5.22 | 0.46 | 0.35 | 4.76 |
| 3 | 5.78 | 0.33 | 0.60 | 5.45 |
| 4 | 13.42 | 0.77 | 1.11 | 12.64 |
| 5 | 21.55 | 0.73 | 0.60 | 20.82 |
| 6 | 8.80 | 0.66 | 1.19 | 8.14 |
| 7 | 3.67 | 1.36 | 0.93 | 2.30 |
| 8 | 10.22 | 2.10 | 2.41 | 8.12 |
| 9 | 6.81 | 2.86 | 1.53 | 3.95 |
| 10 | 18.08 | 2.81 | 3.79 | 15.27 |

**Figure 4.**
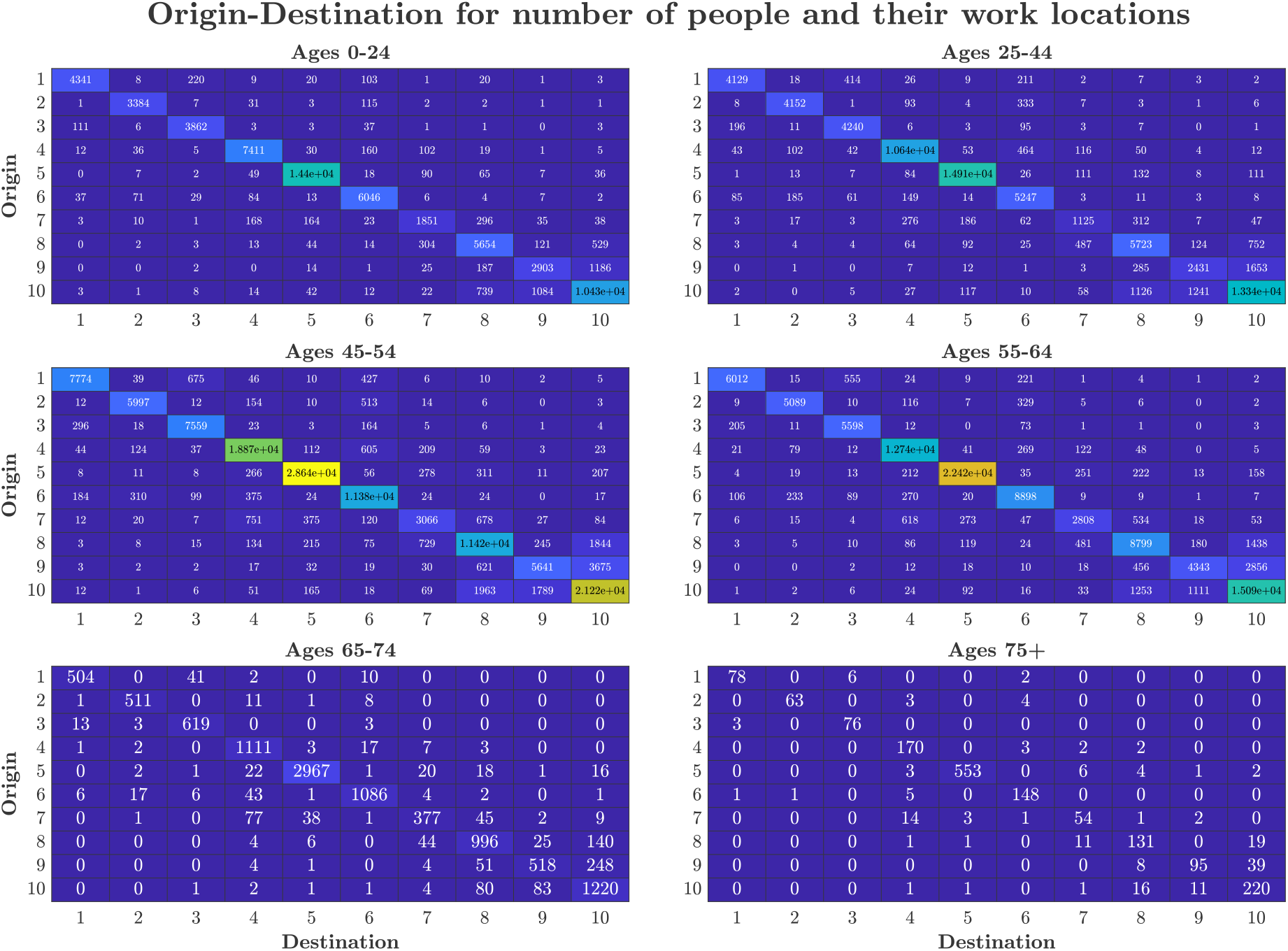
Origin-destination heatmaps illustrating the number of people moving between locations for different age groups. The x-axis represents the destination, and the y-axis represents the origin, with colour intensity and the corresponding numbers indicating the volume of movement.

**Figure 5.**
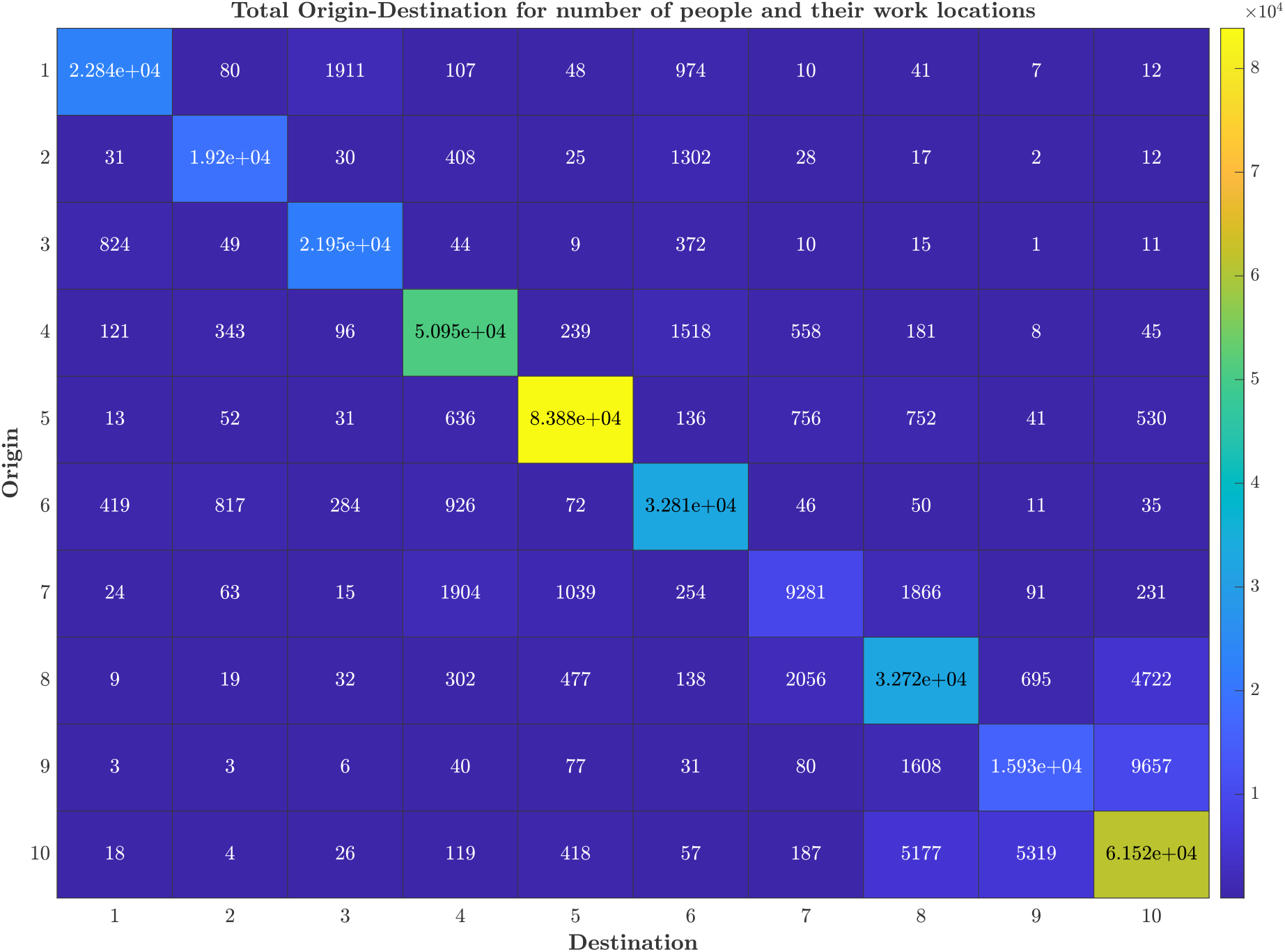
Heatmap of the total origin-destination matrix for all age groups, showing the aggregate number of people moving between locations. The x-axis represents the destination, and the y-axis represents the origin, with colour intensity and the corresponding numbers indicating the volume of movement.

**Figure 6.**
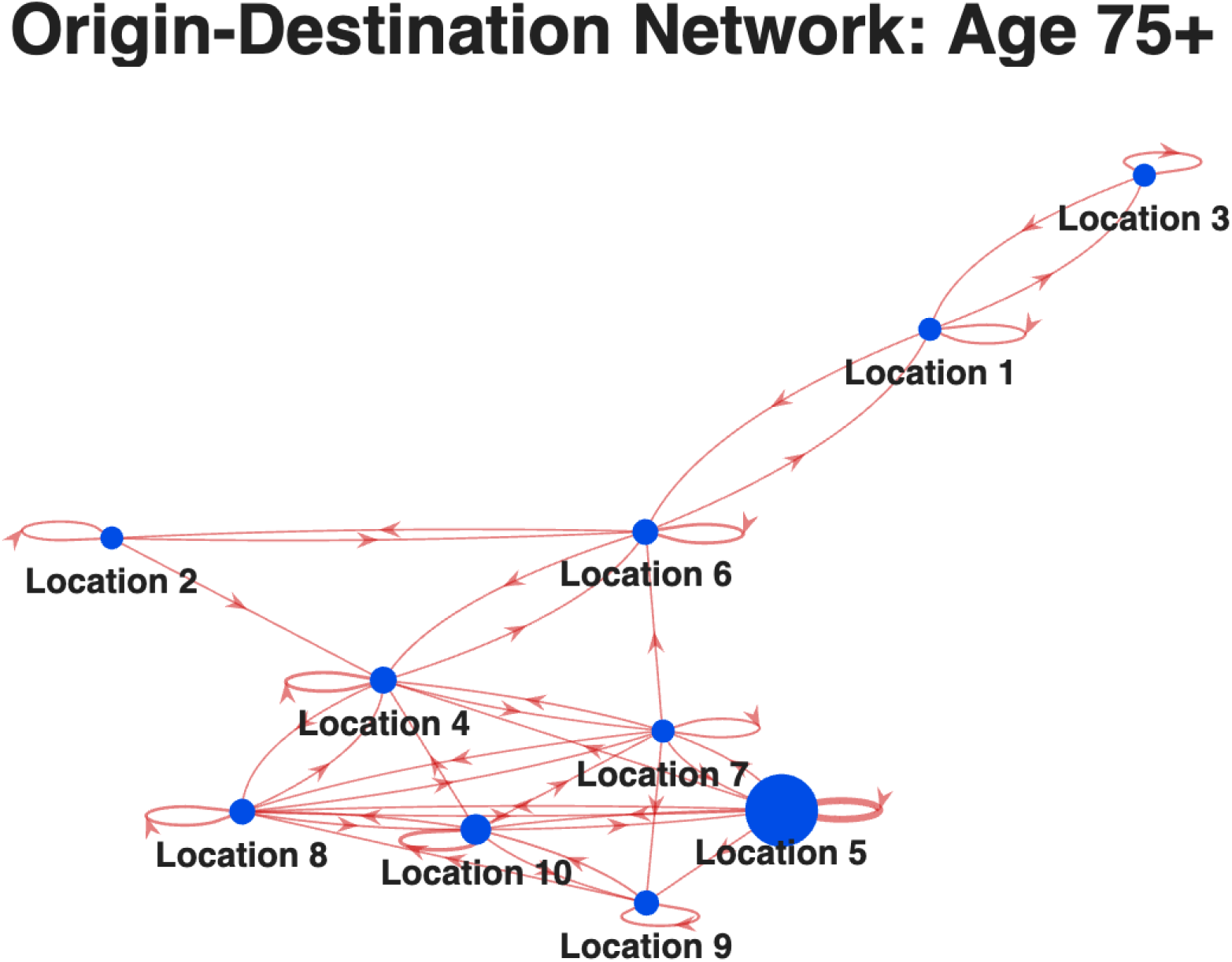
Network visualisation of the origin-destination matrix for the 75+ age group. Nodes represent locations, with sizes proportional to the total outgoing population. Directed edges indicate movement between locations, with weights corresponding to the number of individuals.

**Figure 7.**
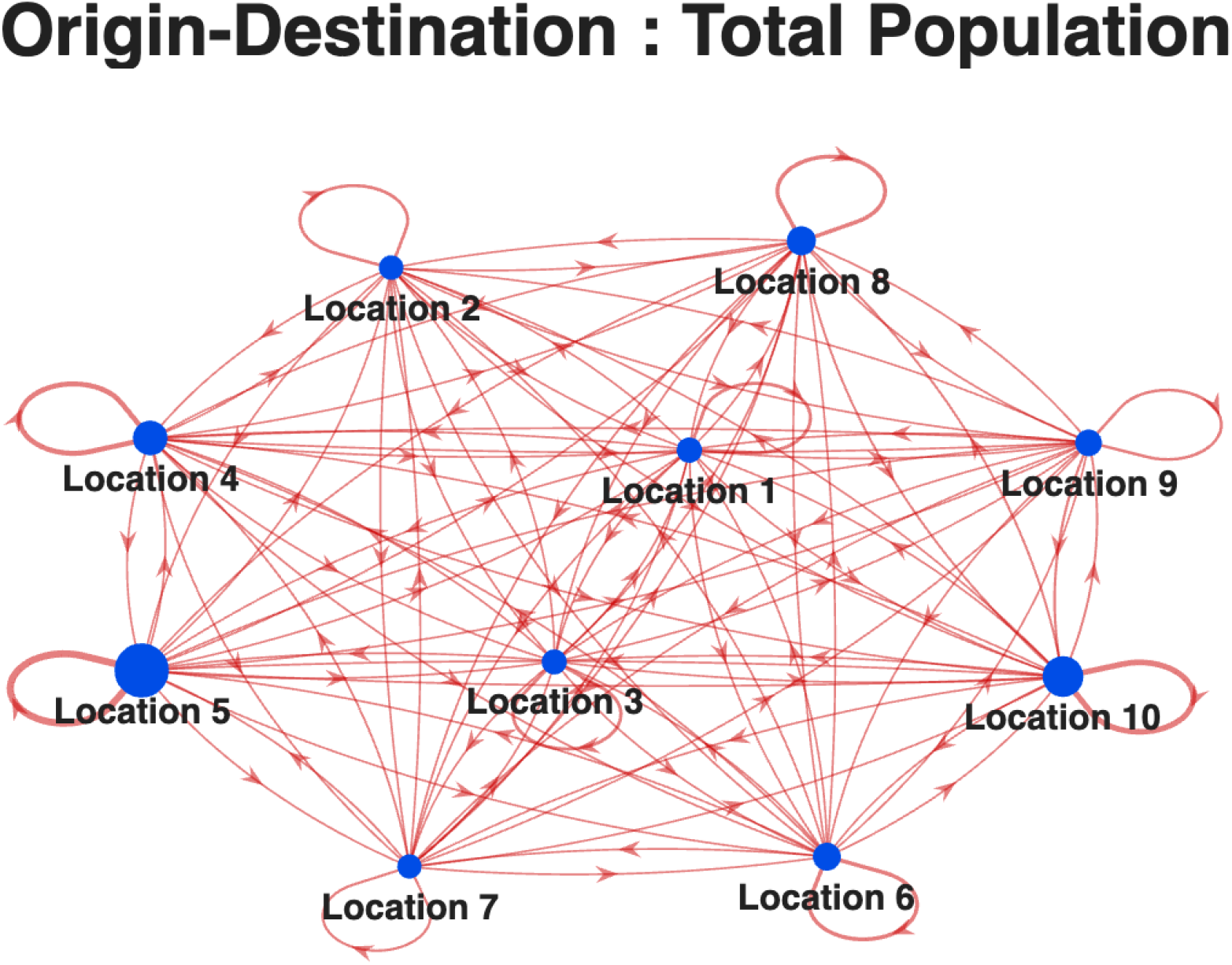
Network visualisation of the total origin-destination matrix across all age groups. Nodes represent locations, with sizes proportional to the total outgoing population. Directed edges indicate movement between locations, with weights corresponding to the number of individuals.

**Figure 8.**
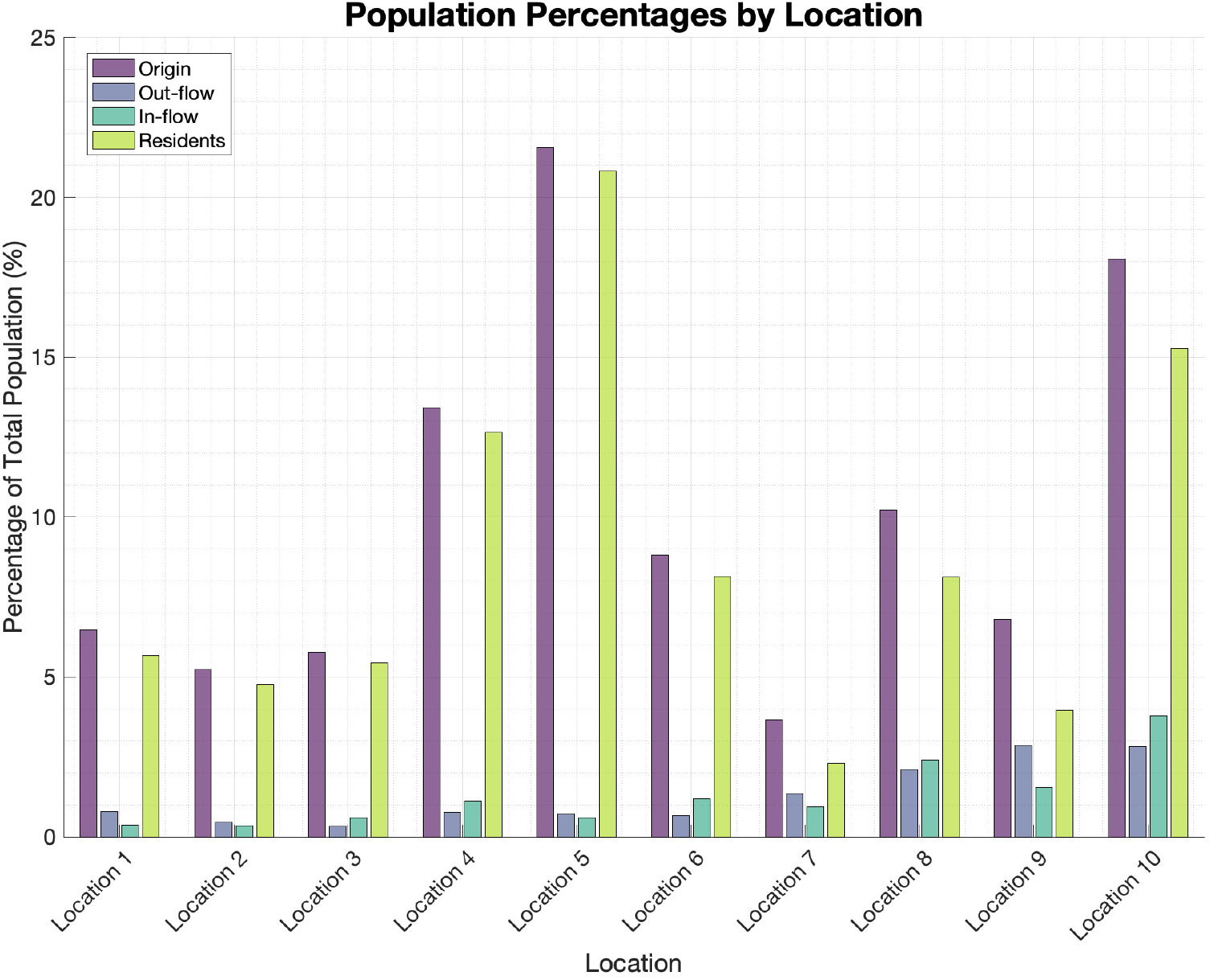
Percentage of total population by location for origin, out-flow, in-flow, and residents, visualised as a grouped bar plot.

#### 2.3.3 COVID-19-like infection parameters

We used the same probability of infection per contact as in the literature [31] as *µ* = 0.5141, the recovery probability per day *γ* = 0.3935, and the probability of progression from exposed to infectious is *σ* = 0.1813. We used the COVID-19 infection hospitalisations risk (IHR) and infection fatality risk (IFR) from the literature [40] as extracted and tabulated in Table 2, and assumed a proportion of asymptomatic (*p*_*a*_) by age groups based on information from the literature [41].

**Table 2.** Infection hospitalisation risk (IHR) and infection fatality risk (IFR) by age for COVID-19 (each value has been divided by 100 to give proportions).

| Age Groups | IHR | IFR |
| --- | --- | --- |
| 6–24 | 0.0036 | 0.000022 |
| 25–44 | 0.0139 | 0.00040 |
| 45–54 | 0.0218 | 0.0020 |
| 55–64 | 0.0434 | 0.0071 |
| 65–74 | 0.0997 | 0.0284 |
| $\geq 75$ | 0.3393 | 0.1695 |

In the numerical simulations of NPIs, we based the initial interaction on the point at which a certain number of cases are reported. Since the level of case reporting can be highly linked to the symptoms of a disease, we factored in the proportion of infections that are expected to result in symptoms as reported cases.

As there was no data available that matched my exact age brackets (6–24, 25–44, 45–54, 55–64, 65–74, 75+), we had to compute the age-dependent proportion of asymptomatic infections for a COVID-19-like infection using other age-specific data. Fortunately, Figure 3 in [41] provided 1-year age estimates for the proportion of asymptomatic infections for COVID-19. This data was estimated from different sample sizes (number infected) across age groups. We assume that the proportion of asymptomatic cases of COVID-19 is reasonably represented by the information in the data and would be useful only for the purpose of demonstrating my model. We used the data for ages 6–90 years, corresponding to the lower age limit in my model and the maximum age in Figure 3 in [41]. Thus, for each age range in my model (e.g., 6–24 years), we average the proportion of asymptomatic infections in the age range as extracted from Figure 3. The summary of the data for the proportion asymptomatic (*p*_*a*_) assumed for the age brackets is tabulated in Table 3.

**Table 3.**
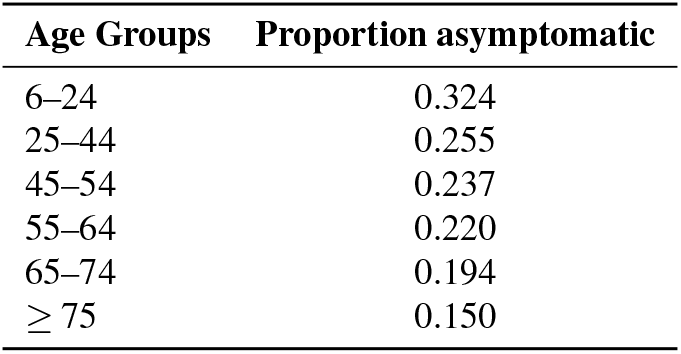
Estimated percentage and proportion of asymptomatic infections by age bracket for a COVID-like infection.

These parameters (IHR, IFR, and *p*_*a*_) are used within my SEIR model to compute the number of hospitalisations, deaths, and symptomatic infections for each age group. For example, using the number of exposed individuals who progress to infectious state at any time, we estimated new hospitalisation that would be expected from this number as 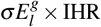, the number of new symptomatic as 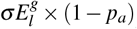, while using the number of removed individuals from the infected population, we computed the number of new deaths as 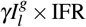. The schematic diagram that depicts this process is in Figure 9.

**Figure 9.**
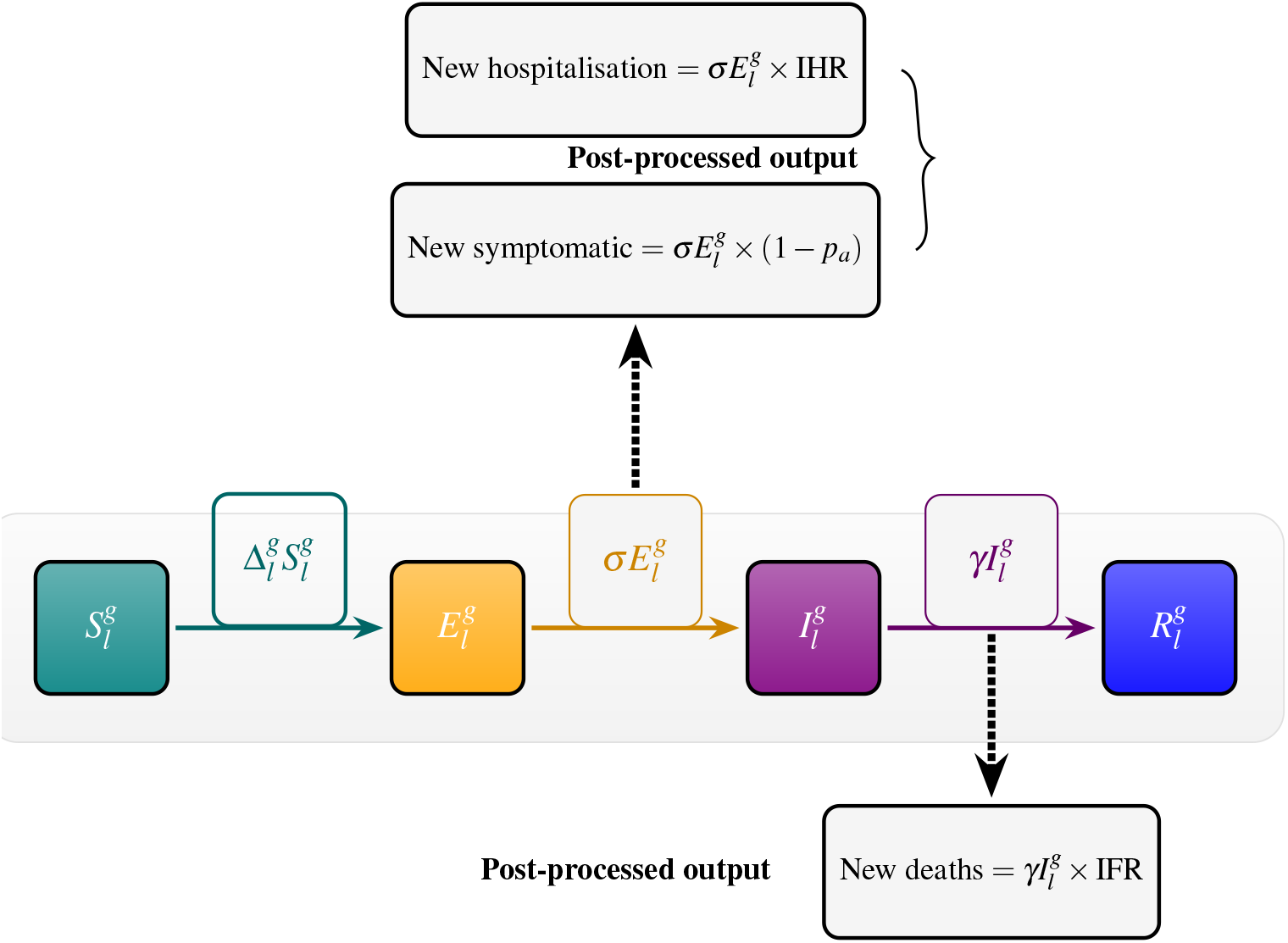
Schematic of the extended location-specific SEIR model for group *g* in location *l*, showing Susceptible 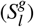, Exposed 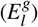, Infectious 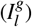, and Recovered 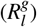 compartments. New hospitalisations and New symptomatic infections are computed using the progression from the exposed, while New deaths are computed from the progression from the infectious. Here, IHR is the infection hospitalisation risk, IFR is the infection fatality risk, and *p*_*a*_ is the proportion asymptomatic.

#### 2.3.4 Ebola-like infection parameters

Ebola is transmitted by contact (it is not an airborne disease) and is generally considered contagious when there are symptoms [42]. Based on this, we have made a simplifying assumption that there are no asymptomatic infections. The incubation and infectious period are 9.4 and 7.8 days, respectively [43]. Since contagiousness/infectiousness comes with symptoms, we assumed that the latency period (the time between when a exposed and when a person become infectious) is the same as the incubation period (the time between being exposed to infection and when symptoms first appear). Hence, the rate of progression from being exposed to infectious and the recovery rate are 1*/*9.4 days^−1^ and 1*/*7.8 days^−1^ respectively. We converted the rates to probabilities to have the recovery probability *γ* = 0.1203, and the progression probability, *σ* = 0.1009. Using the basic reproduction number of ℛ_0_ = 1.5 [43], and the rest of the parameter values, we calculated the value of the probability of per contact infection for Ebola using a method similar to that in the literature [31] to *µ* = 0.0873. We utilised the case fatality risk (CFR) for Ebola in the literature [44] and assume that it would represent the IFR, with asymptomatic infection uncommon [45]. To match the CFR with age stratification in my model, we used CFR data for ages 5 − 15 years (average CFR for ages 5 − 6, 7 − 9, 10 − 15) to represent the IFR for ages 6 − 24 in my model, we use CFR for ages 16-44 to represent the IFR for ages 25 − 44 and CFR for ages 44+ to represent the IFR for ages 45 − 54, 55 − 64, 65 − 74 and 75+ in my model. We assume the infection hospitalisation risk to be 80%. This is within the range of the percentage of healthcare-seeking individuals in the literature [43]. Because this data was not available for the age groups in my model, we generalised this IHR across all age groups. The IFR and IHR that we used for an Ebola-like infection are summarised in Table 4.

**Table 4.** Assumed infection hospitalisation risk (IHR) and infection fatality risk (IFR) by age for an Ebola-like infection.

| Age Groups | IHR | IFR |
| --- | --- | --- |
| 6–24 | 0.80 | 0.588 |
| 25–44 | 0.80 | 0.646 |
| 45–54 | 0.80 | 0.768 |
| 55–64 | 0.80 | 0.768 |
| 65–74 | 0.80 | 0.768 |
| $\geq 75$ | 0.80 | 0.768 |

### 2.4 Numerical simulation

We implemented numerical simulations of epidemic outbreaks in three scenarios. Initially, we introduced the first infection in Location 1 by setting the number of initially infected individuals to zero in all other locations in the metapopulation except for Location 1. Then, we simulated the outbreak and applied different interventions. This process was repeated with the initial infection starting in Location 5, and finally, in Location 10. Among these three locations (as detailed in Table 1 and Figure 8), Location 5 has the highest population size, accounting for 21.55% of the total population, followed by Location 10 (18.08%), and Location 1 (6.46%). Location 10 has the highest number of visitors (in-flow), accounting for 3.79% of the total population visiting for work/school, and gives the highest number of travellers (out-flow) to other locations, accounting for 2.81% of the total population travelling from this location to work/school. Location 5 has the highest number of its residents (Residents) going to work/school within their location, accounting for 20.82% of the total population who travel to work/school in their original locations, while Location 10’s Residents account for 15.27%, Location 1 has the lowest resident percentage (5.67%), consistent with its lower origin. In all cases, the number of people who travel to work within their locations of residence supersedes their inflows and outflows. Summarily, Location 5 is a dominant origin and resident hub with little movement, Location 1 is a small, stable area, and Location 10 shows substantial travel typical of a commercial city. Seeding initial infection in each of these locations would allow me to deduce the impact of the location of initial infections on the overall outbreak trajectories in the presence/absence of interventions.

For each of the locations of initial infection, we simulated three intervention strategies;

1. No intervention (“No NPI”),
2. Total work/school closure (“Total closure”) in all locations for all age groups when a total of ten (symptomatic) infections have been reported,
3. Targeted work/school closure (“Targeted closure”) in affected locations for all age groups when a total of ten (symptomatic) infections have been reported.

When the reported infection threshold to react (ten symptomatic infections across all locations) is met, we allow a three-day delay before implementing intervention. This is to allow for the possibility of reaction delay to allow people to prepare to adjust (for example, public health planning and messaging, and the population to buy foodstuffs) before the onset of interventions. In the case of the targeted closure, It is possible that one or two locations may contain exposed individuals but not symptomatic (reported) infections at the time when the threshold of ten reported cases across the whole population is met. Although such location(s) may eventually report cases before the start of intervention, the declaration of targeted intervention (which should have occurred earlier) leaves them out. This is to account for the effect of the latency period and proportion of asymptomatic infection in a targeted intervention, as locations that are considered not affected and left out of intervention plans may actually have either asymptomatic infections or exposed individuals who may contribute to new outbreaks in those locations.

Numerical simulations are implemented for both deterministic (Model (12)) and stochastic (Model (16)) to compare their results. Except for mobility probability, we assume statistical equivalence for the same age group across all locations, so that the parameter values for each age group apply in all locations.

To compute the probability of a large outbreak (defined here as at least three locations recording at least 20 infections each), we count the number of realisations that meet this criterion and divide by the total number of realisations that we implemented. For all stochastic results presented in this study (both COVID-like and Ebola-like), we run 1,000 realisations.

## 3 Results

### 3.1 COVID-19-like infection

#### 3.1.1 Deterministic vs stochastic model

Initially, we compared epidemic trajectories for COVID-19-like infection, produced by deterministic and stochastic simulations across three NPI strategies and different seeding locations (Location 1, 5, or 10) (Figure 10). In the absence of NPIs, both stochastic and deterministic simulations show qualitatively similar curves, as seen by rapid epidemic growth and a single peak. However, deterministic models consistently produced higher epidemic peaks compared to the mean of stochastic simulations. Stochastic realisations (grey lines) show visible variability in outbreak size and timing, including the possibility of early outbreak fade-out, which deterministic models cannot capture.

**Figure 10.**
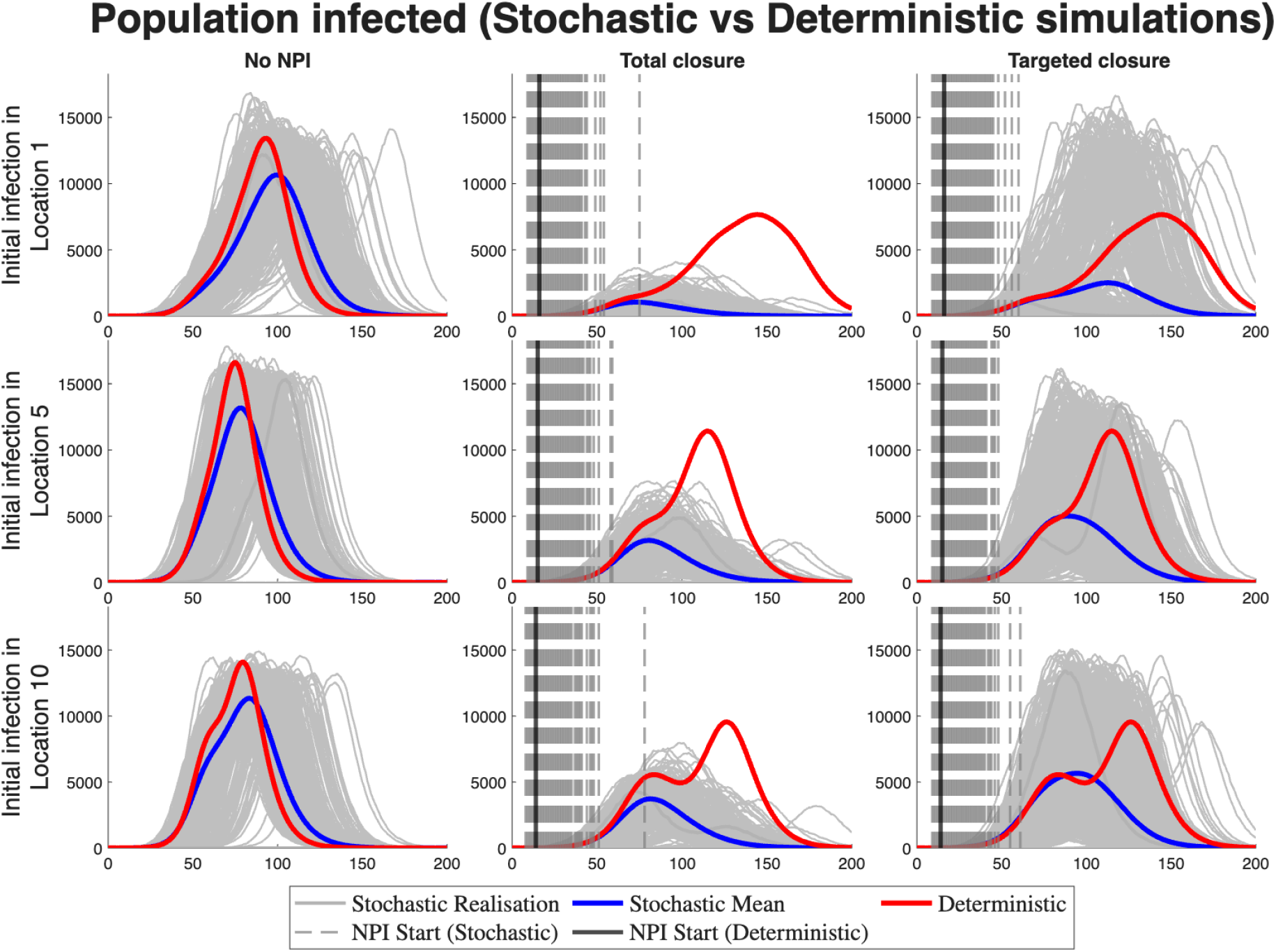
COVID-19-like infection: Comparison of total population infected in stochastic versus deterministic simulations across different initial locations of infection scenarios and intervention strategies. The first row is the results of starting the initial infection in age group 6 − 24 in Location 1, the second row is the results of starting the initial infection in age group 6 − 24 in Location 5, and the third row is the results of starting the initial infection in age group 6 − 24 in Location 10. The first column of each row is the case without intervention, the second column is the total work/school closure, while the third column is the total work/school closure in affected locations at the time of intervention. The grey bars are the timing of NPI starts for all the different stochastic realisations, while the black solid bar is the timing of starting NPI for the deterministic model.

“Total closure of schools and workplaces across all locations (middle column, Figure 10) effectively flattened the epidemic curve and delayed peaks. Deterministic simulations under this strategy show a secondary peak, whereas stochastic simulations indicate a slower epidemic growth and a greater likelihood of control. The targeted closure of schools and workplaces in affected areas (right column) shows reduced peaks compared to the no intervention strategy, but allows for greater spread than complete closure. In all scenarios, deterministic models show visibly higher outbreak size with multiple peaks. The reason for this variability is that, in stochastic simulations, locations without infected, exposed, or asymptomatic individuals at the beginning of interventions remain uninfected, whereas for the deterministic simulation, less than one but greater than zero infected person is possible, hence after interventions are implemented, the fractional infected person grows leading to eventual outbreaks in locations with less than one infected person at the beginning of intervention. Again, this emphasises how the stochastic approach captures more realistic outbreak scenarios than their deterministic counterparts. Therefore, for later plots and tables, we only consider the outputs from the stochastic model.

Seeding infections in different locations (rows of Figure 10) shows that, in the absence of NPIs (the first column of Figure 10), the infection would almost certainly take off. The overall shape, timing, and epidemic peaks are comparable for initial infection in Locations 5 and 10, with differences falling within the variability of stochastic realisations. The initial seeding infection in Location 1 results in a slightly late outbreak and lower peak. This suggests that once transmission is established, the epidemic trajectory is largely independent of the initial location of introduction due to mixing between locations. However, there are visible differences in infection trajectories for different locations of the initial outbreak when interventions are implemented, with the highest peak at Location 10, followed by Location 5, when the total closure is implemented and the highest peak at Location 5, followed by Location 10, when the targeted closure is implemented. This variability highlights the role of the size and the connectivity of the initial location in the overall outcome of the epidemic, which includes the design and implementation of interventions.

#### 3.1.2 Age- and location-specific patterns

We analysed the age-specific and location-specific epidemic under NPIs for COVID-19–like infections. The infections (mean of stochastic realisation) by location in Figure 11 and the proportion of each age group that got infected (Figure 12) show that without NPIs, all locations eventually experience outbreaks, although these show different peak distributions depending on the location of initial infection. Under the total closure, infections often remain localised or fade out entirely, whereas the targeted closure slow but do not prevent multi-location spread.

**Figure 11.**
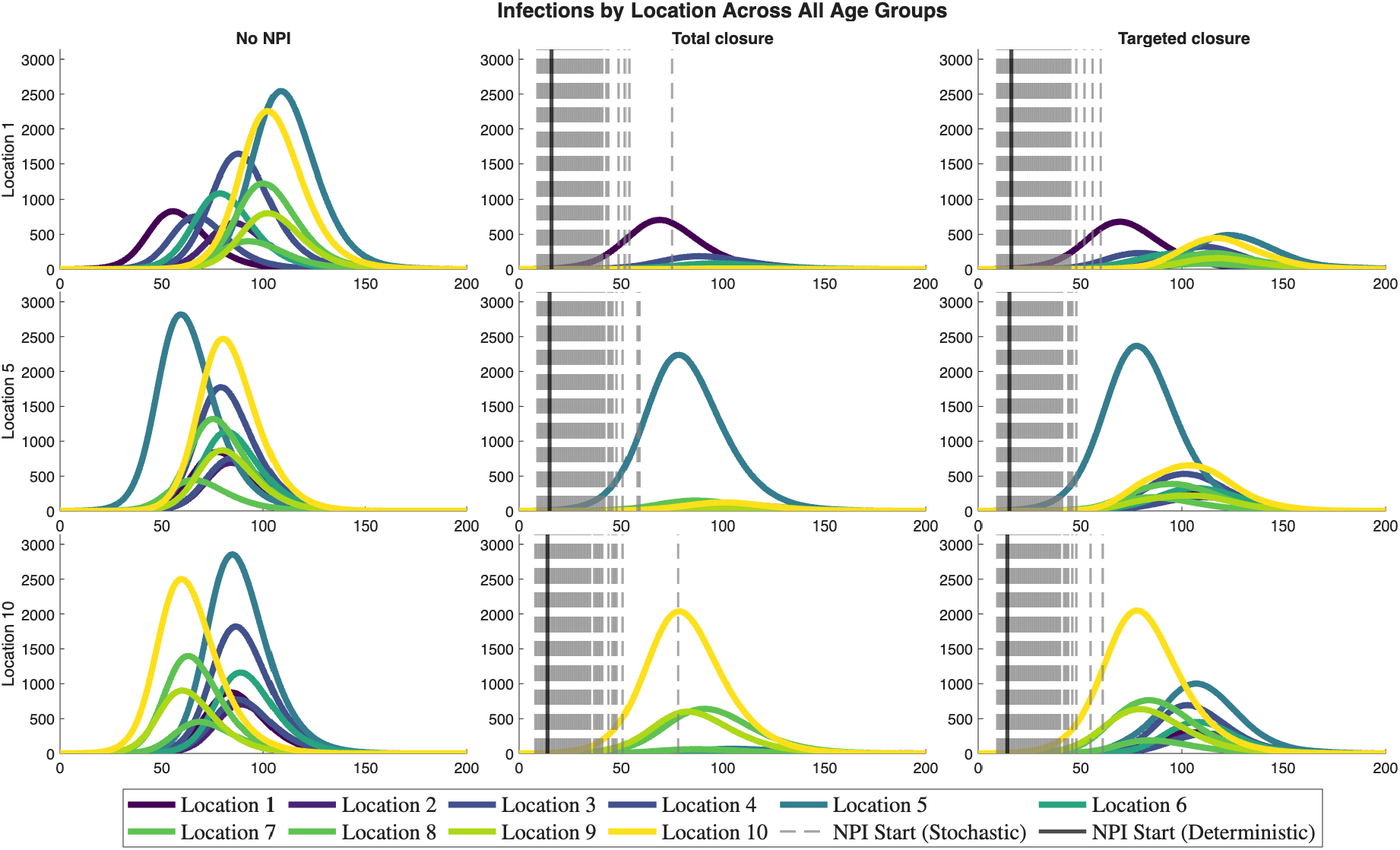
COVID-19-like infection: Mean stochastic infections by location across all age groups for different scenarios and intervention strategies. The first row is the results of starting the initial infection in age group 6 − 24 in Location 1, the second row is the results of starting the initial infection in age group 6 − 24 in Location 5, and the third row is the results of starting the initial infection in age group 6 − 24 in Location 10. The first column of each row is the case without intervention, the second column is the total work/school closure, while the third column is the total work/school closure in affected locations at the time of intervention. The grey bars are the timing of NPI starts for all the different stochastic realisations, while the black solid bar is the timing of starting NPI for the deterministic model.

**Figure 12.**
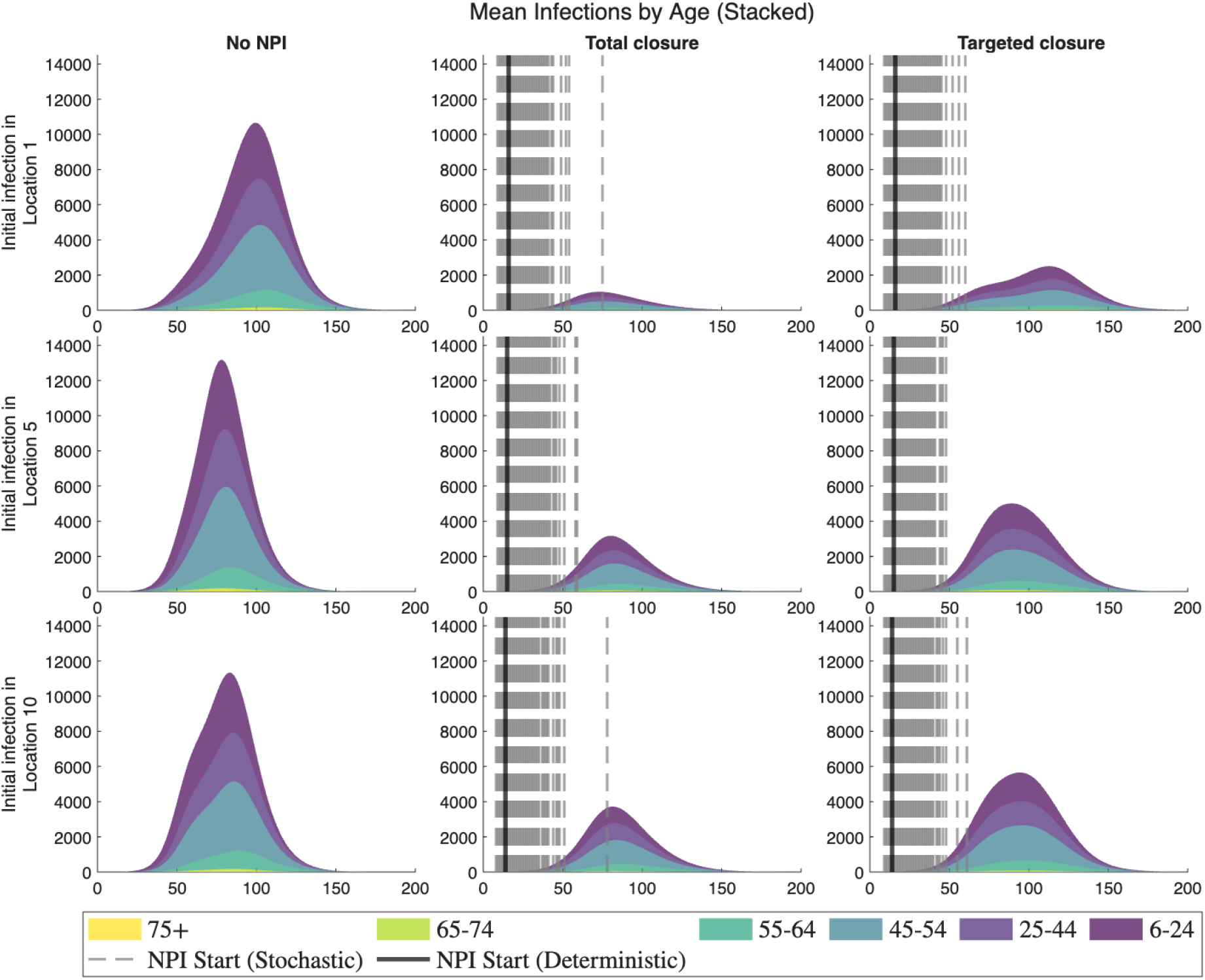
COVID-19-like infection: Mean number of individuals infected in each age group across all realisations, shown as stacked area plots. The first row is the results of starting the initial infection in age group 6 − 24 in Location 1, the second row is the results of starting the initial infection in age group 6 − 24 in Location 5, and the third row is the results of starting the initial infection in age group 6 − 24 in Location 10. The first column of each row is the case without intervention, the second column is the total work/school closure, while the third column is the total work/school closure in affected locations at the time of intervention. The grey bars are the timing of NPI starts for all the different stochastic realisations, while the black solid bar is the timing of starting NPI for the deterministic model.

**Figure 13.**
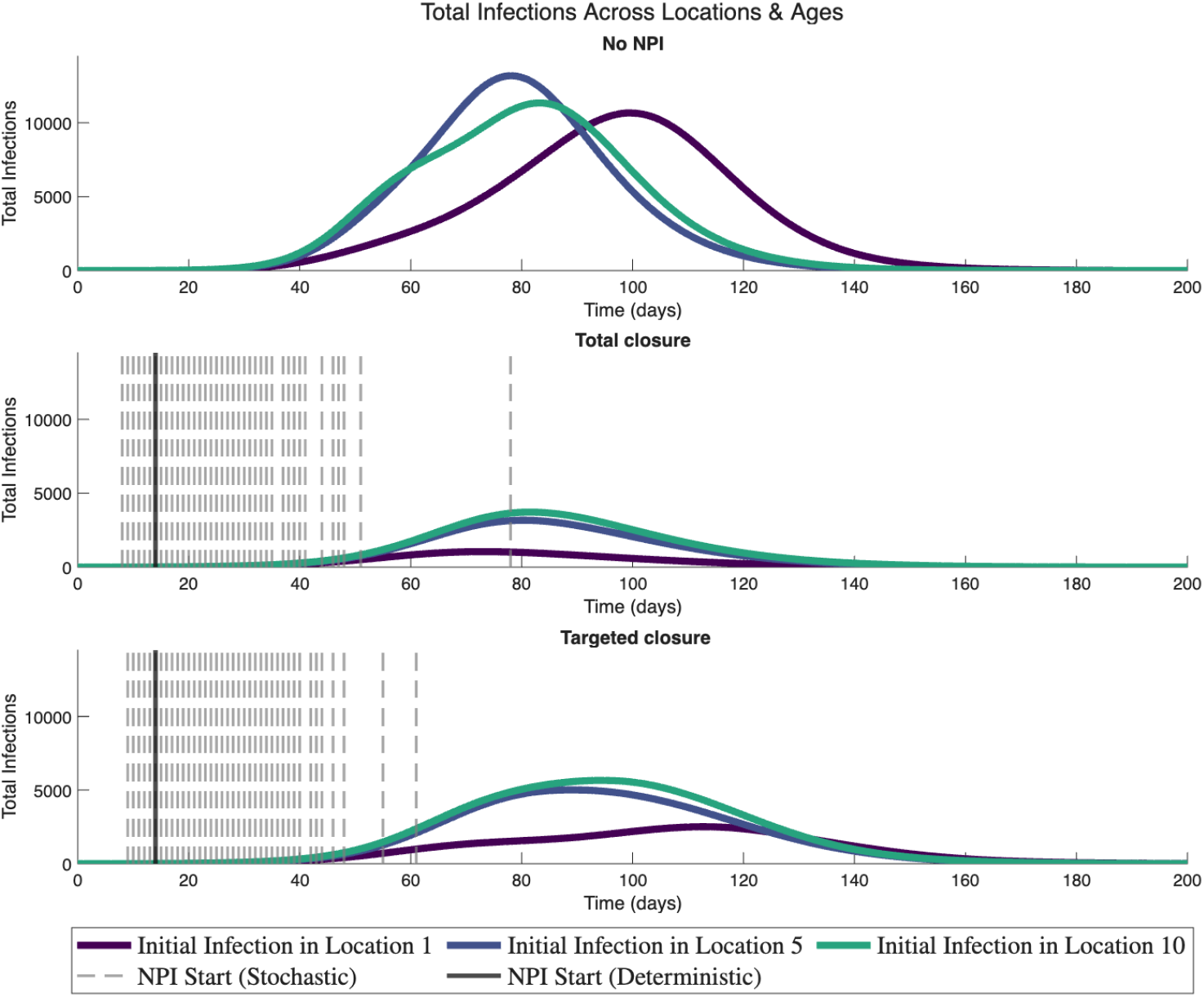
COVID-19-like infection: Infections across all locations and age groups for different scenarios and intervention strategies. The grey bars are the timing of NPI starts for all the different stochastic realisations, while the black solid bar is the timing of starting NPI for the deterministic model.

The final epidemic size distributions (Figure 14) also reveal contrasts between scenarios. The “No NPIs” consistently show large outbreaks with little variation. Under interventions, especially the total closures, many epidemics fade out early, while others proceed to smaller but still non-negligible sizes. These fade-outs are absent from deterministic simulations, emphasising the importance of stochastic approaches for policy-relevant insights.

**Figure 14.**
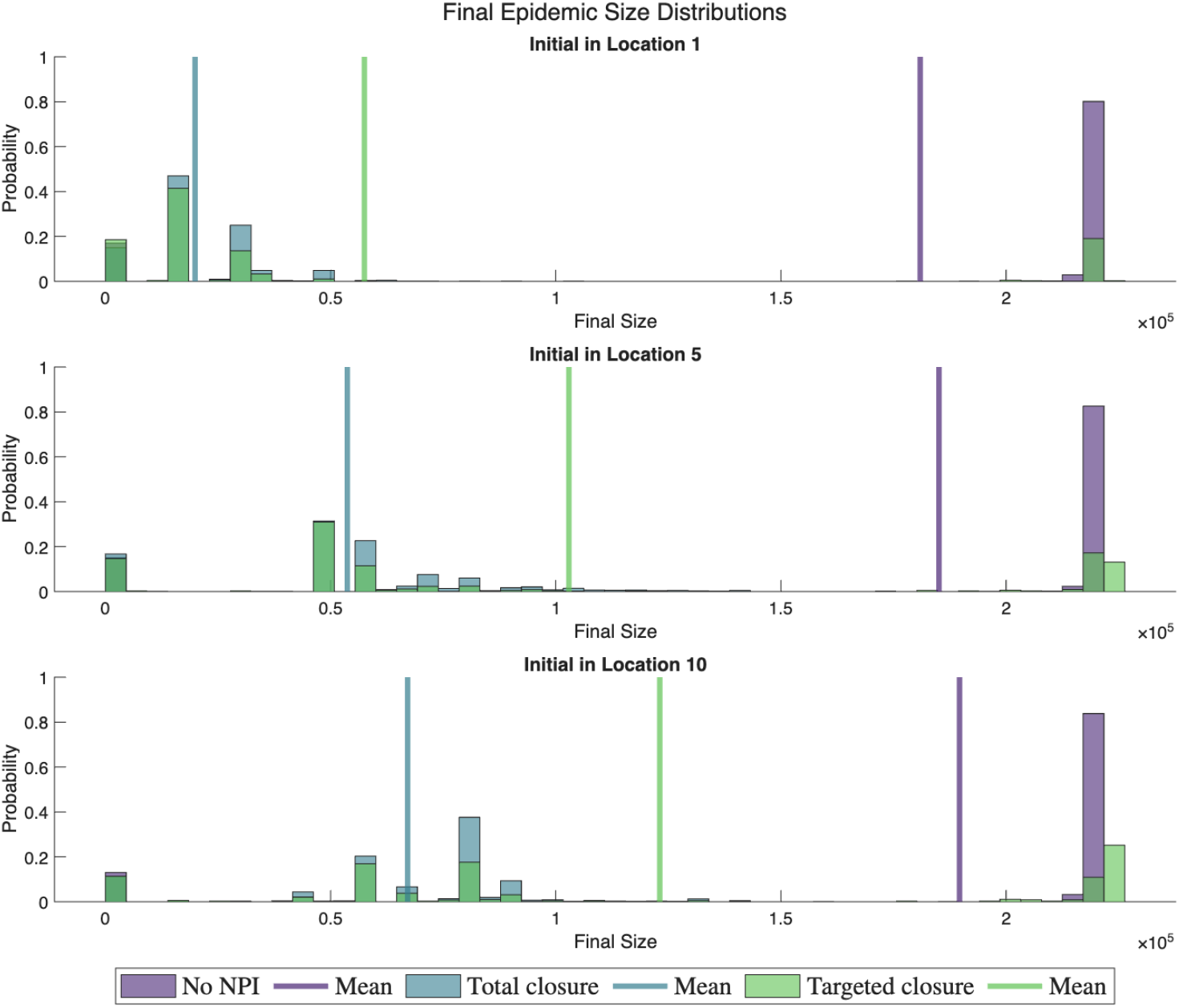
COVID-19-like infection: Final epidemic size distributions across all scenarios, shown as histograms for different intervention strategies.

The cumulative impacts are reflected in total hospitalisations and deaths (Figure 15). Without interventions, both metrics remain very high across all seeding locations. The total closures produce the highest reductions in both hospitalisations and deaths, followed by the targeted closure. The benefits of NPIs are consistent across seeding scenarios but strongest when outbreaks begin in Location 1. This is due to the limited inflow and outflow of people, including the small population size in Location 1.

**Figure 15.**
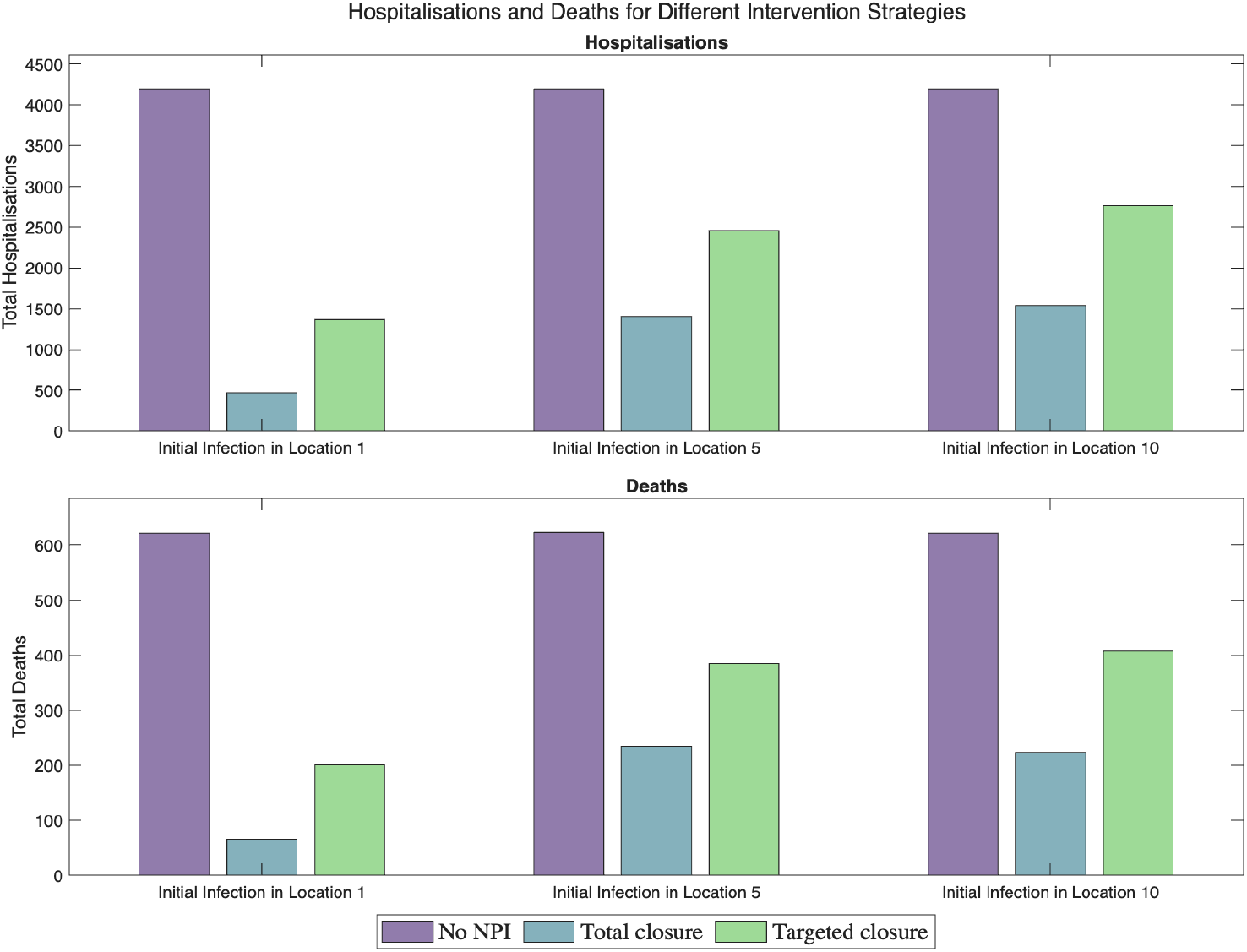
COVID-19-like infection: Total hospitalisations and deaths for different intervention strategies, shown as grouped bar plots.

Daily hospitalisations by age group (Figure 16) show that without NPIs, sharp epidemic peaks emerge across all seeding locations scenarios. The total closure reduces and delays hospitalisations across all age groups, followed by the targeted closure in affected locations.

**Figure 16.**
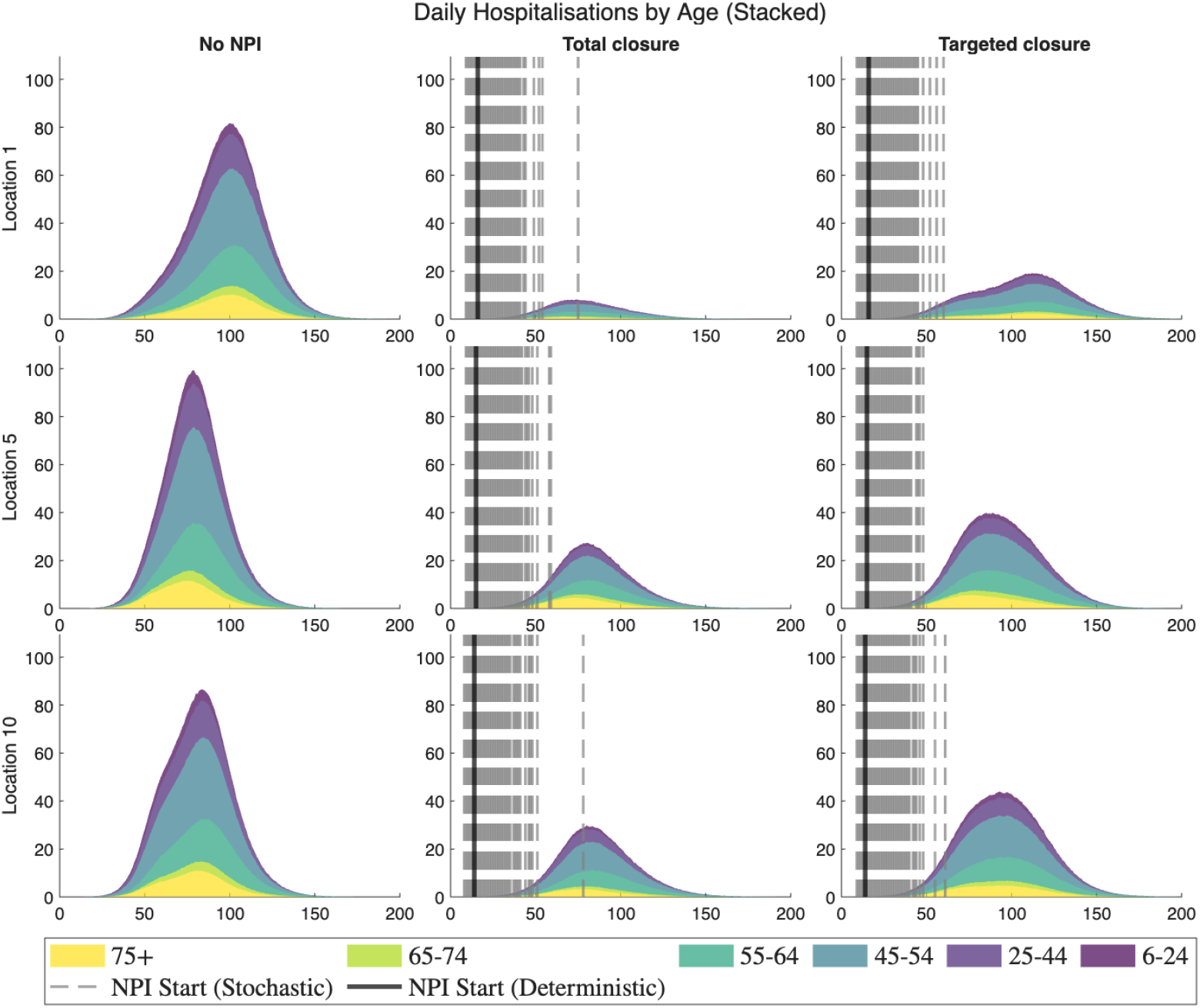
COVID-19-like infection: Daily hospitalisations by age group for different scenarios and intervention strategies, shown as stacked area plots. The first row is the results of starting the initial infection in age group 6 − 24 in Location 1, the second row is the results of starting the initial infection in age group 6 − 24 in Location 5, and the third row is the results of starting the initial infection in age group 6 − 24 in Location 10. The first column of each row is the case without intervention, the second column is the total work/school closure, while the third column is the total work/school closure in affected locations at the time of intervention. The grey bars are the timing of NPI starts for all the different stochastic realisations, while the black solid bar is the timing of starting NPI for the deterministic model.

The total daily hospitalisations in all locations and age groups (Figure 17) further illustrate that the total closures effectively flatten the peaks, essentially reducing the peak hospital burden and easing the demand for healthcare. The targeted closure also achieve a reduction in the peak but allow substantial outbreaks.

**Figure 17.**
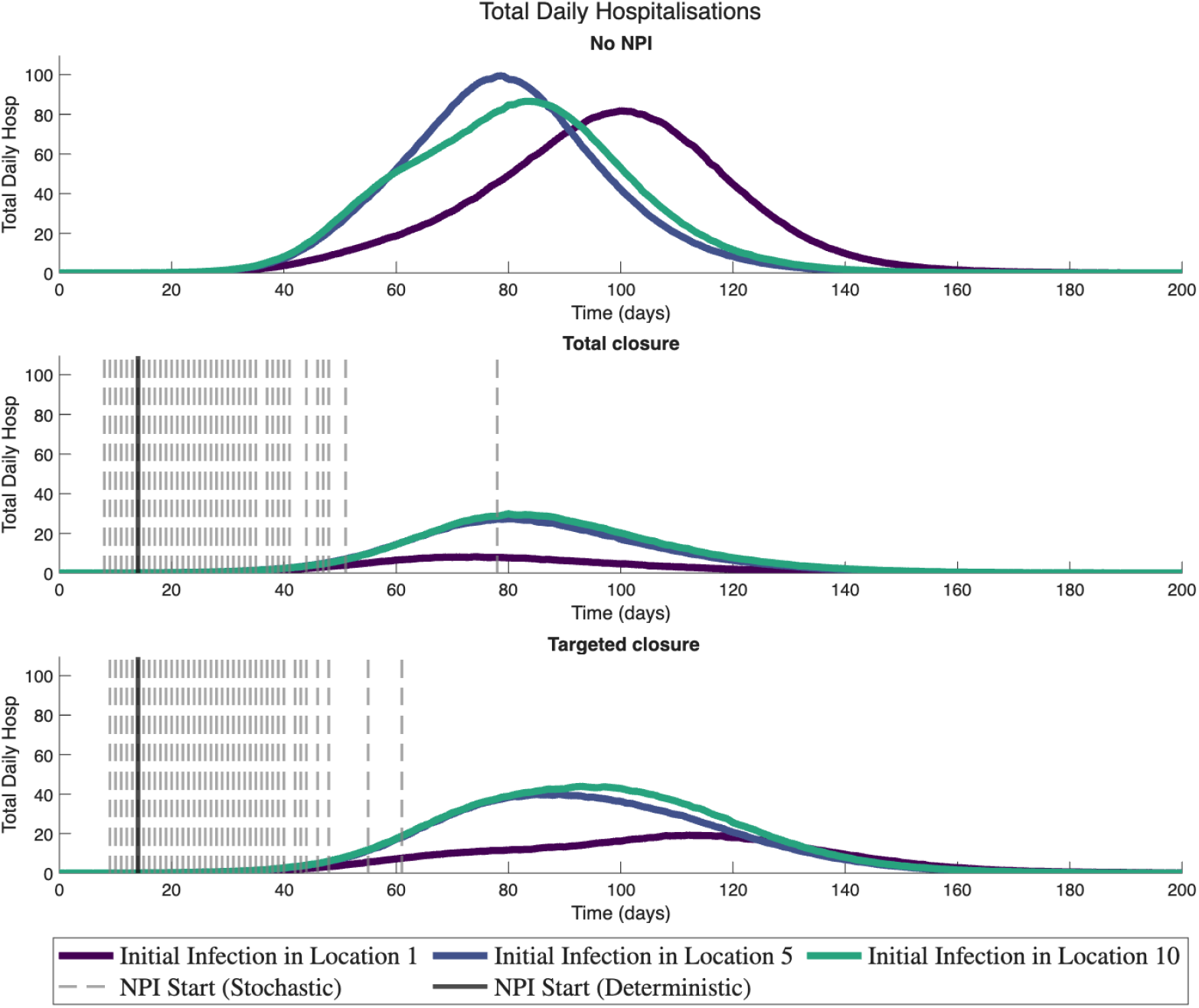
COVID-19-like infection: Daily hospitalisations across all locations and age groups for different scenarios and intervention strategies. The grey bars are the timing of NPI starts for all the different stochastic realisations, while the black solid bar is the timing of starting NPI for the deterministic model.

Figures 13, 17, and 18 show that the highest and earliest epidemic peak results from initial infection in Location 5, particularly in the absence of NPIs; initial infection in Location 10 dominates in the higher and earliest peaks when interventions are implemented. For the infected by age group (Figures 19 and 20), the highest peaks are observed for ages 6–64 when initial infection is seeded in Location 10, but the highest peaks in ages 65 and above for initial infection seeded in Location 5.

**Figure 18.**
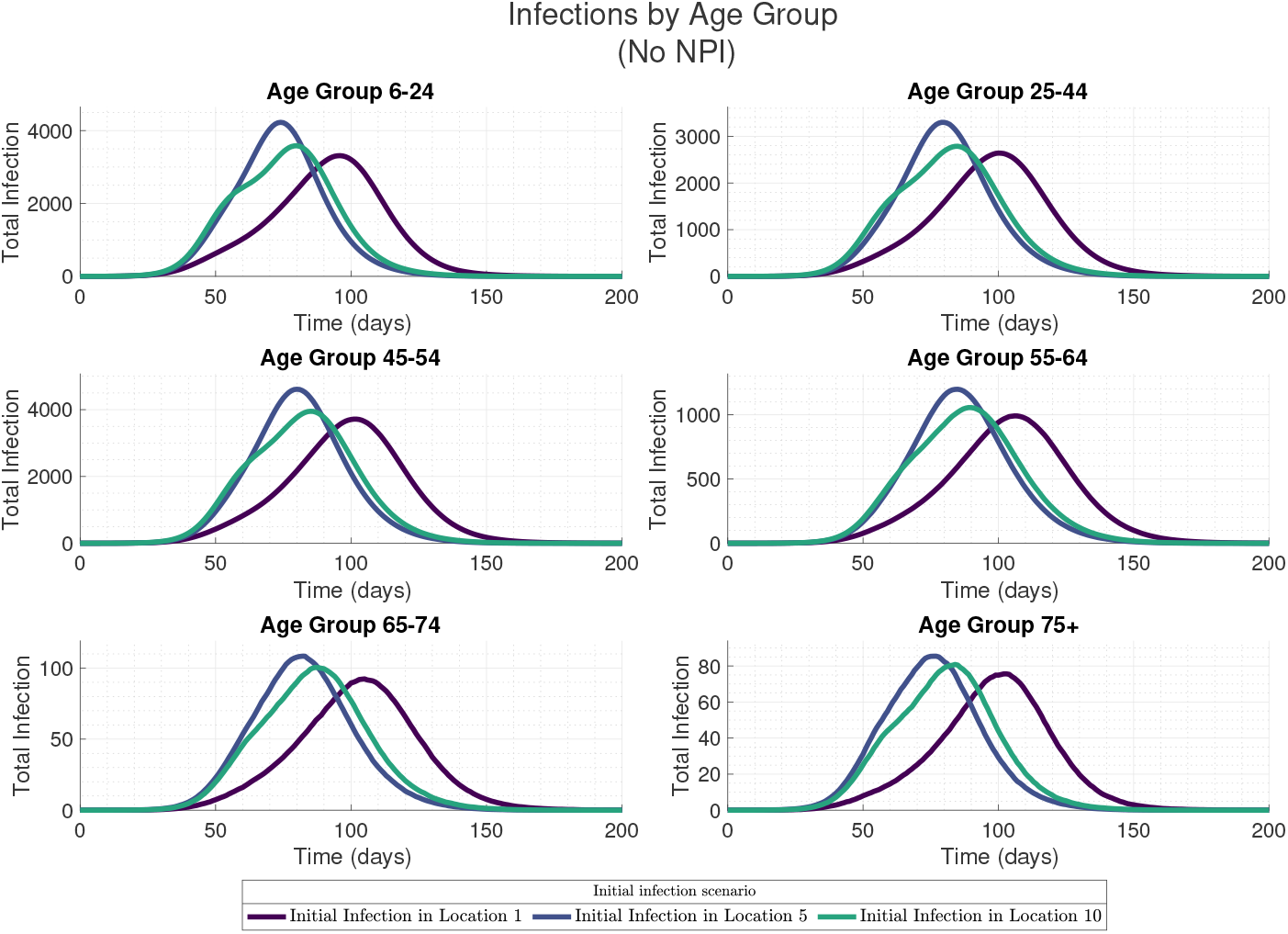
COVID-19-like infection: Infections for each age group across all locations with no interventions.

**Figure 19.**
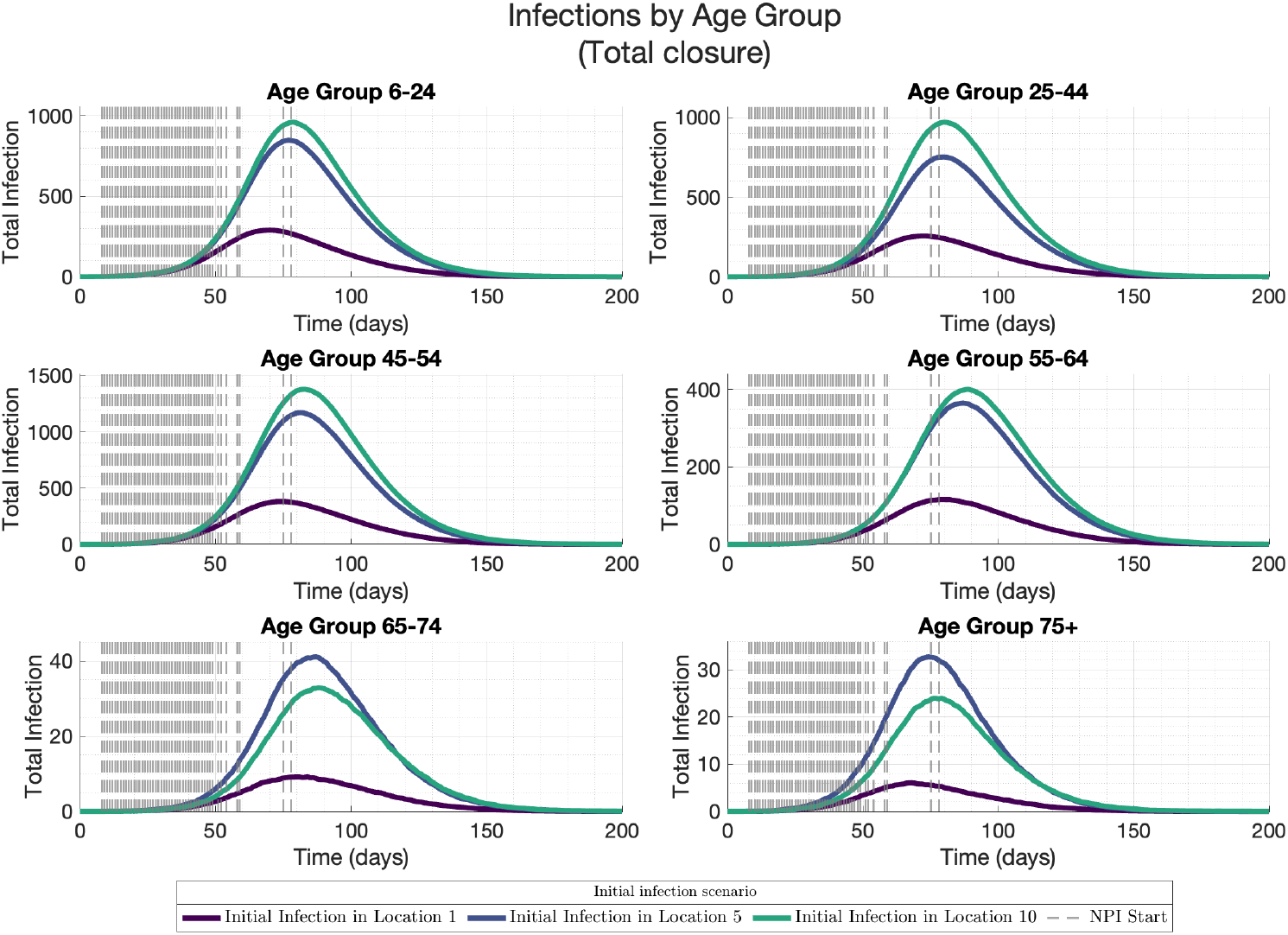
COVID-19-like infection: Infections for each age group across all locations with work/school closure in all locations.

**Figure 20.**
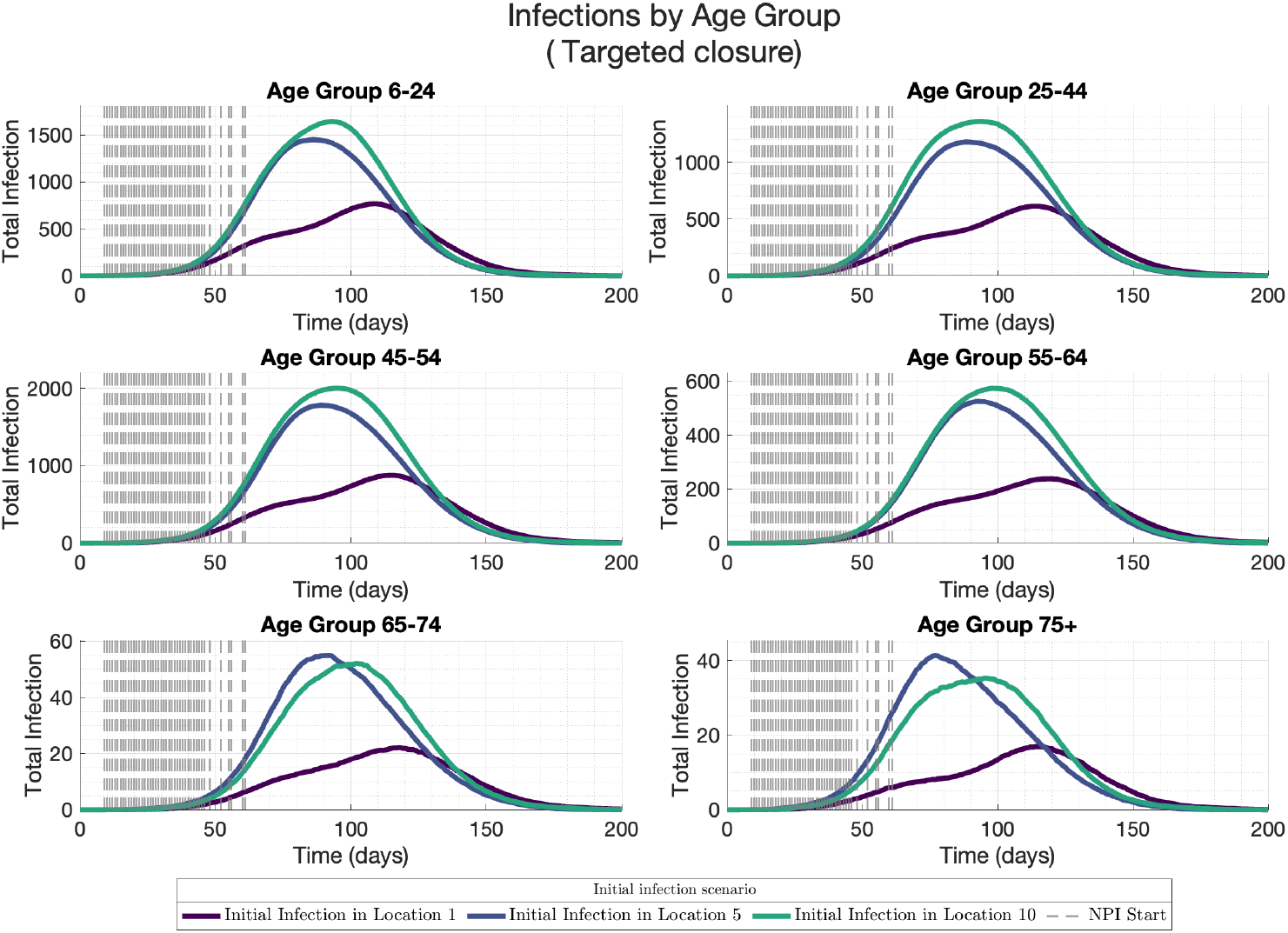
COVID-19-like infection: Infections for each age group across all locations with work/school closure in affected locations.

### 3.2 Ebola-like infection

#### 3.2.1 Deterministic vs stochastic model

Similar to the deterministic and stochastic simulations across three NPI strategies and different seeding locations (Location 1, 5, or 10) for the COVID-19-like infection, in the absence of NPIs, both stochastic and deterministic simulations of the Ebola-like infection show qualitatively similar curves, as seen in Figure 21.

**Figure 21.**
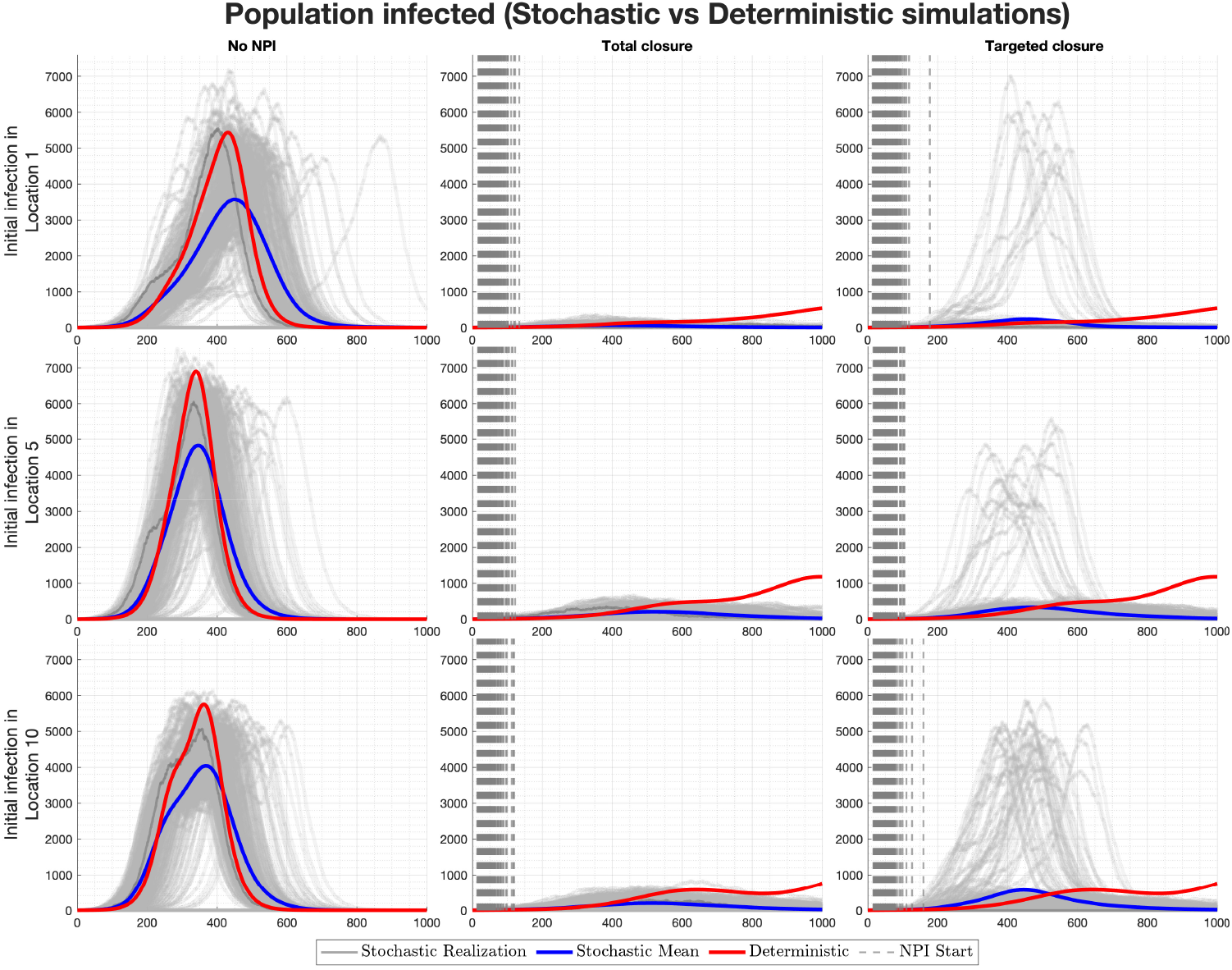
Ebola-like infection: Comparison of total population infected in stochastic versus deterministic simulations across different initial locations of infection scenarios and intervention strategies. The first row is the results of starting the initial infection in age group 6 − 24 in Location 1, the second row is the results of starting the initial infection in age group 6 − 24 in Location 5, and the third row is the results of starting the initial infection in age group 6 − 24 in Location 10. The first column of each row is the case without intervention, the second column is the total work/school closure, while the third column is the total work/school closure in affected locations at the time of intervention. The grey bars are the timing of NPI starts for all the different stochastic realisations, while the black solid bar is the timing of starting NPI for the deterministic model.

The total closure of schools and workplaces across all locations (middle column, Figure 21) effectively flattened the epidemic curve. Deterministic simulations under this strategy show a divergence from the mean of stochastic realisations. The targeted closure of schools and workplaces in affected areas (right column) shows reduced peaks compared to the no intervention strategy, but allows for greater spread than complete closure. This approach shows some major outbreaks for the stochastic realisation, which are not captured using the deterministic simulations. Again, this emphasises how the stochastic approach captures more realistic outbreak scenarios than their deterministic counterparts.

Similar to the COVID-19-like infection in Figure 10, seeding infections in different locations (rows of Figure 21) shows that, in the absence of NPIs (the first column of Figure 21), the infection would almost certainly take off. The peak infection is higher when initial infection is seeded in Location 5, followed by Location 10, and Location 1 respectively. The timing are comparable for initial initial infection in Locations 5 and 10, with differences falling within the variability of stochastic realisations. However, seeding initial infection in Location 1 results in a little late outbreak. The stochastic realisations show that for the targeted closure, there are few major outbreaks for infection seeded in any of the three locations.

#### 3.2.2 Age- and location-specific patterns

The mean stochastic infections by location in Figure 22 show that without NPIs, all locations eventually experience outbreaks, although these show different peak distributions depending on the location of initial infection. Under the total closure and the targeted closure, infections remain localised or fade out entirely.

**Figure 22.**
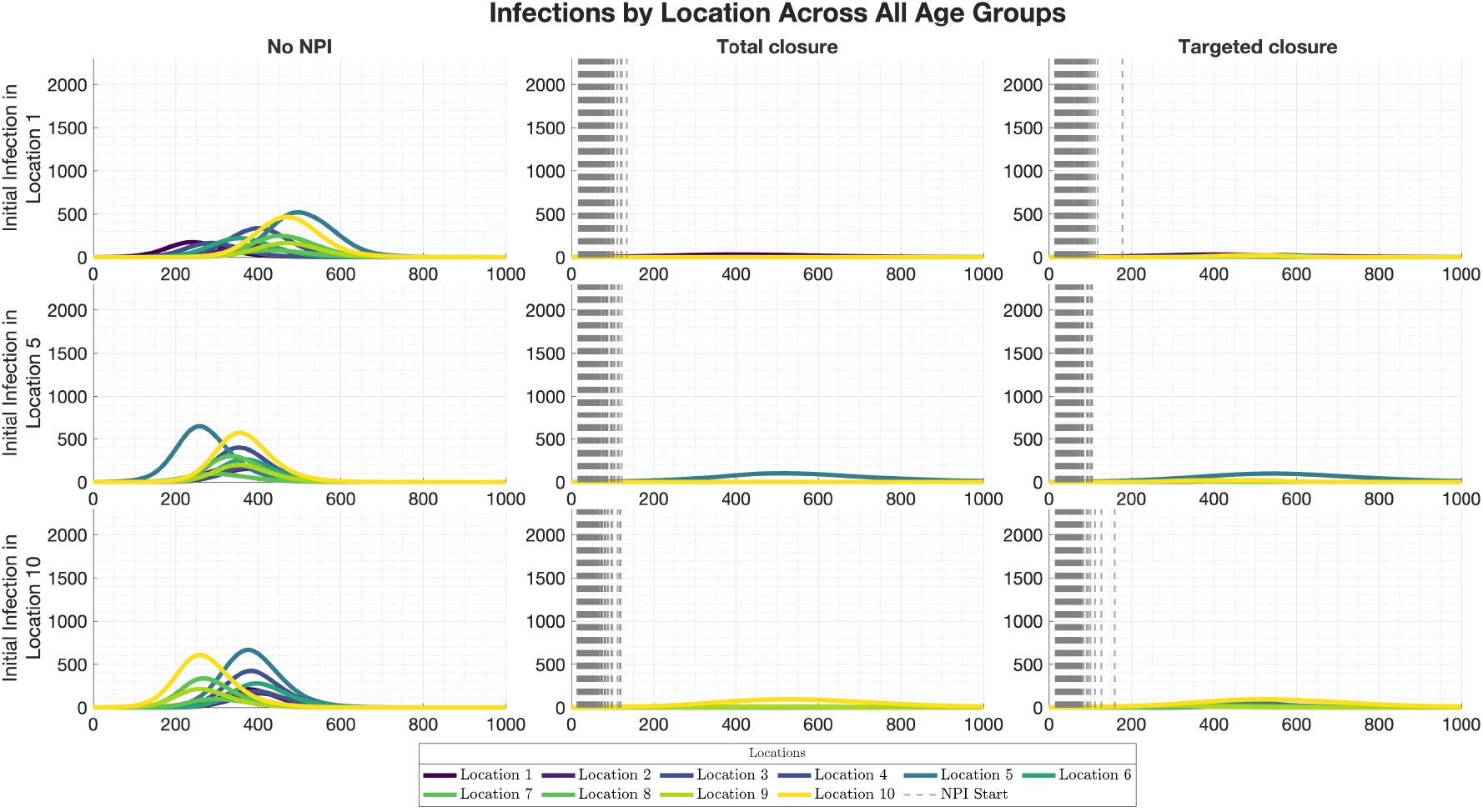
Ebola-like infection: Mean stochastic infections by location across all age groups for different scenarios and intervention strategies. The first row is the results of starting the initial infection in age group 6 − 24 in Location 1, the second row is the results of starting the initial infection in age group 6 − 24 in Location 5, and the third row is the results of starting the initial infection in age group 6 − 24 in Location 10. The first column of each row is the case without intervention, the second column is the total work/school closure, while the third column is the total work/school closure in affected locations at the time of intervention.

### 3.3 Comparing COVID-like and Ebola-like infections

Comparisons between COVID-19–like (Figures 10-20) and Ebola-like infections (Figures 21-31) reveal the strong influence of pathogen characteristics on epidemic outcomes and intervention effectiveness. COVID-19–like epidemics grew rapidly and reached high peaks, even under interventions, whereas Ebola-like epidemics spread more slowly, producing delayed and lower peaks. The absence of asymptomatic infections in the Ebola-like simulation presented here may have influenced the slower spread and the effectiveness of NPIs, as NPIs are more effective in the Ebola-like setting, often preventing large outbreaks altogether. These contrasts highlight how pathogen-specific transmissibility and infectious period shape both epidemic trajectories and the effectiveness of interventions.

**Figure 23.**
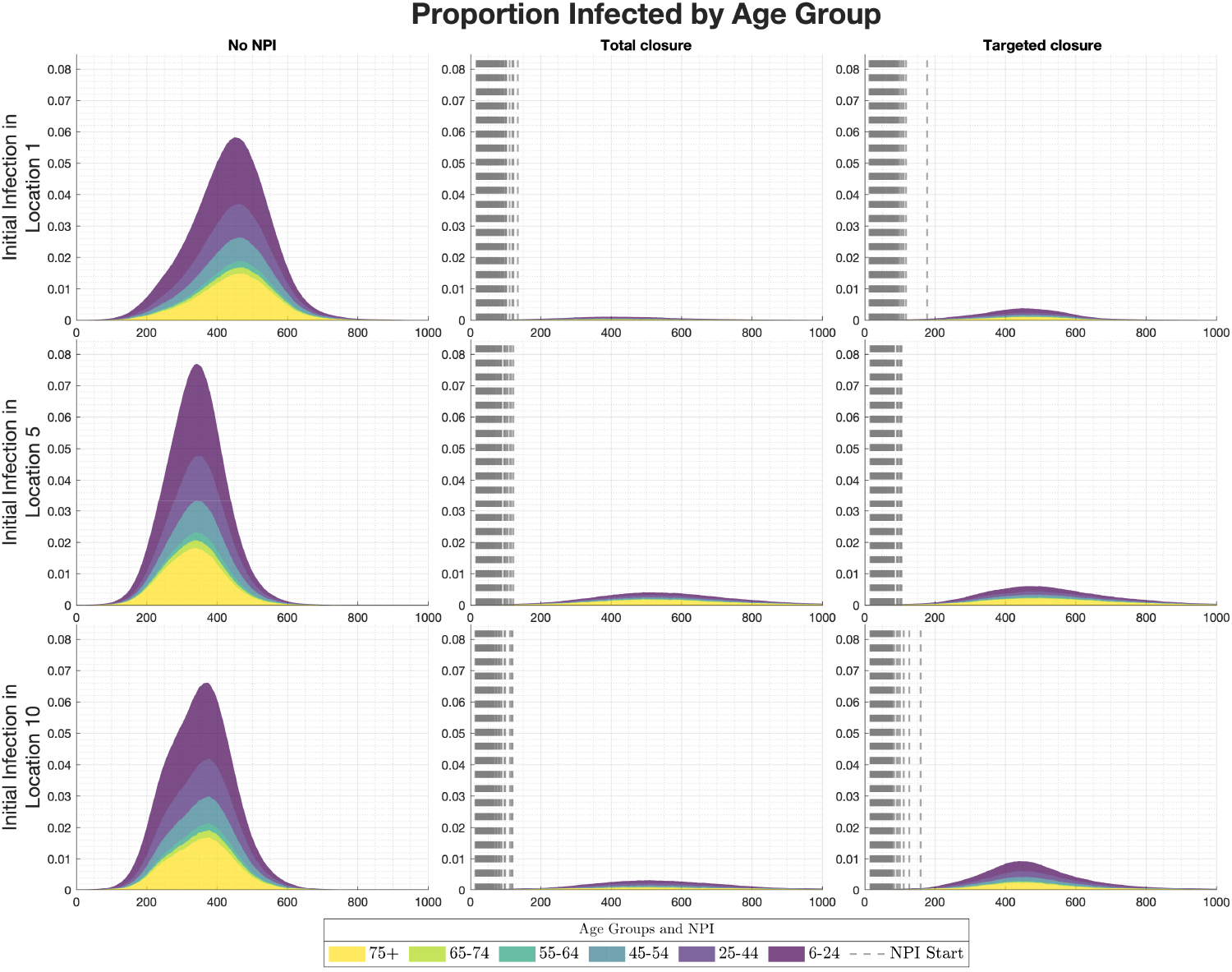
Ebola-like infection: Mean of the proportion of each age group that got infected for different scenarios and intervention strategies across all realisations, shown as stacked area plots. The first row is the results of starting the initial infection in age group 6 − 24 in Location 1, the second row is the results of starting the initial infection in age group 6 − 24 in Location 5, and the third row is the results of starting the initial infection in age group 6 − 24 in Location 10. The first column of each row is the case without intervention, the second column is the total work/school closure, while the third column is the total work/school closure in affected locations at the time of intervention.

**Figure 24.**
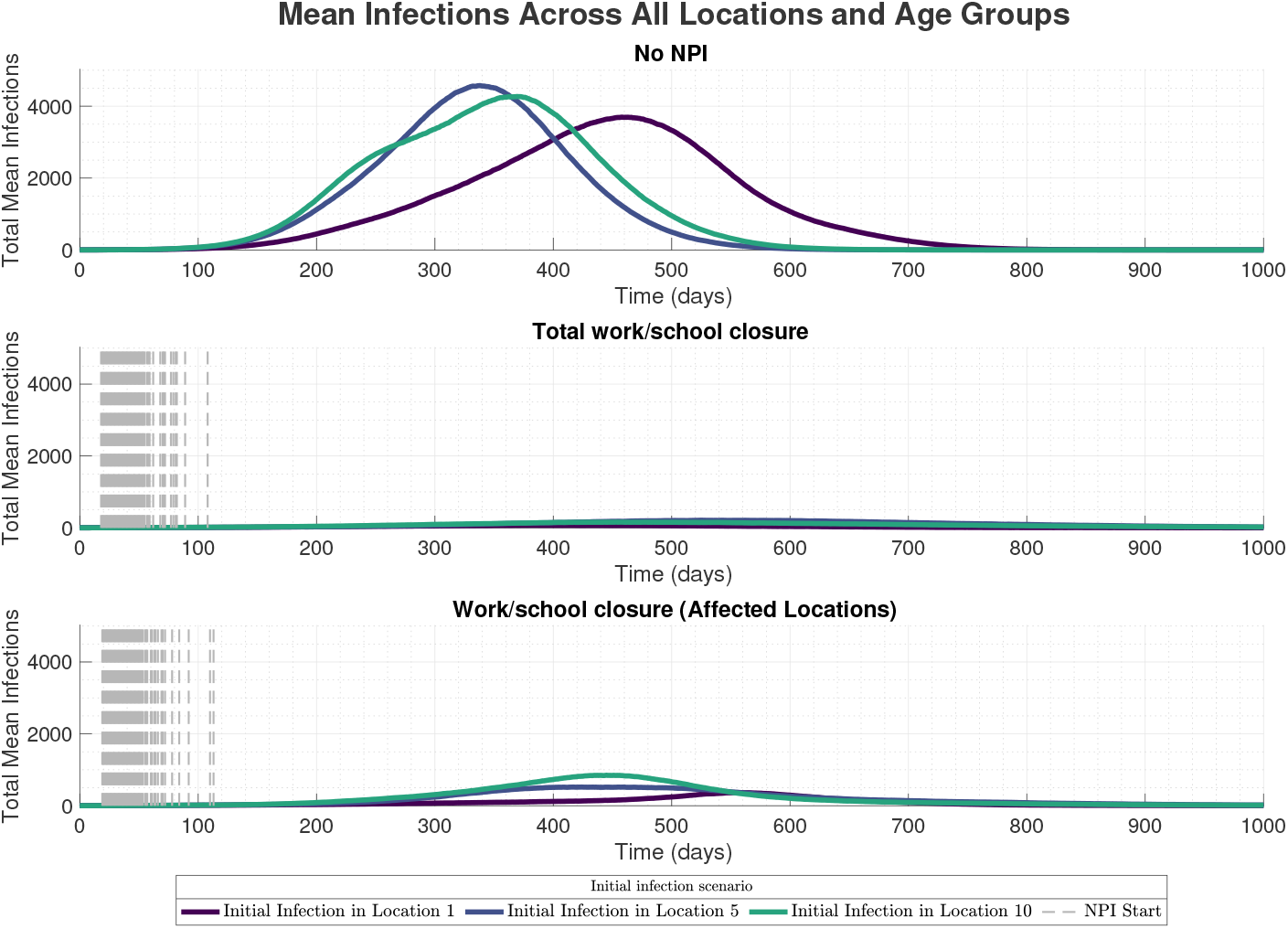
Ebola-like infection: Infections across all locations and age groups for different scenarios and intervention strategies.

**Figure 25.**
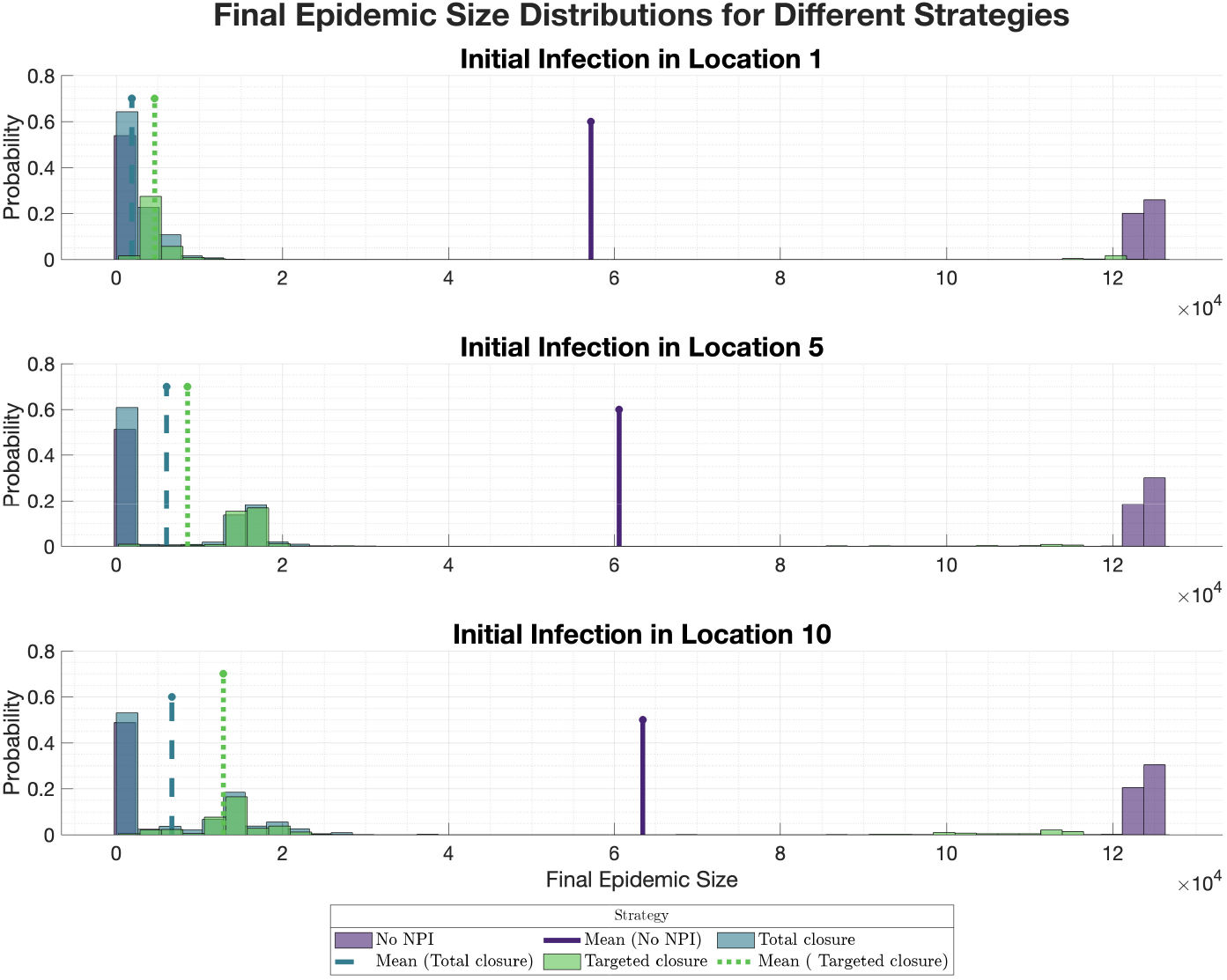
Ebola-like infection: Final epidemic size distributions across all scenarios, shown as histograms for different intervention strategies.

**Figure 26.**
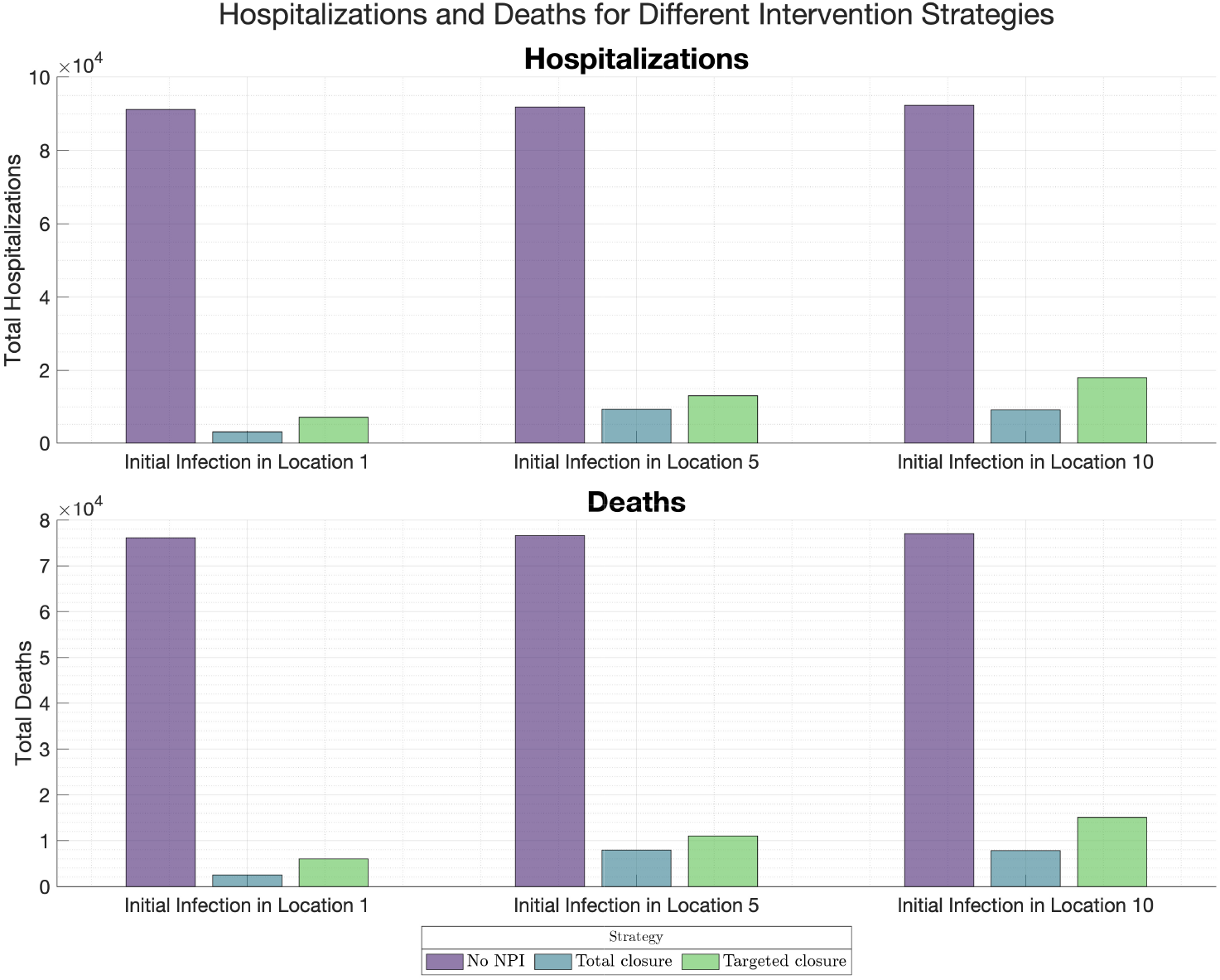
Ebola-like infection: Total hospitalisations and deaths for different intervention strategies, shown as grouped bar plots.

**Figure 27.**
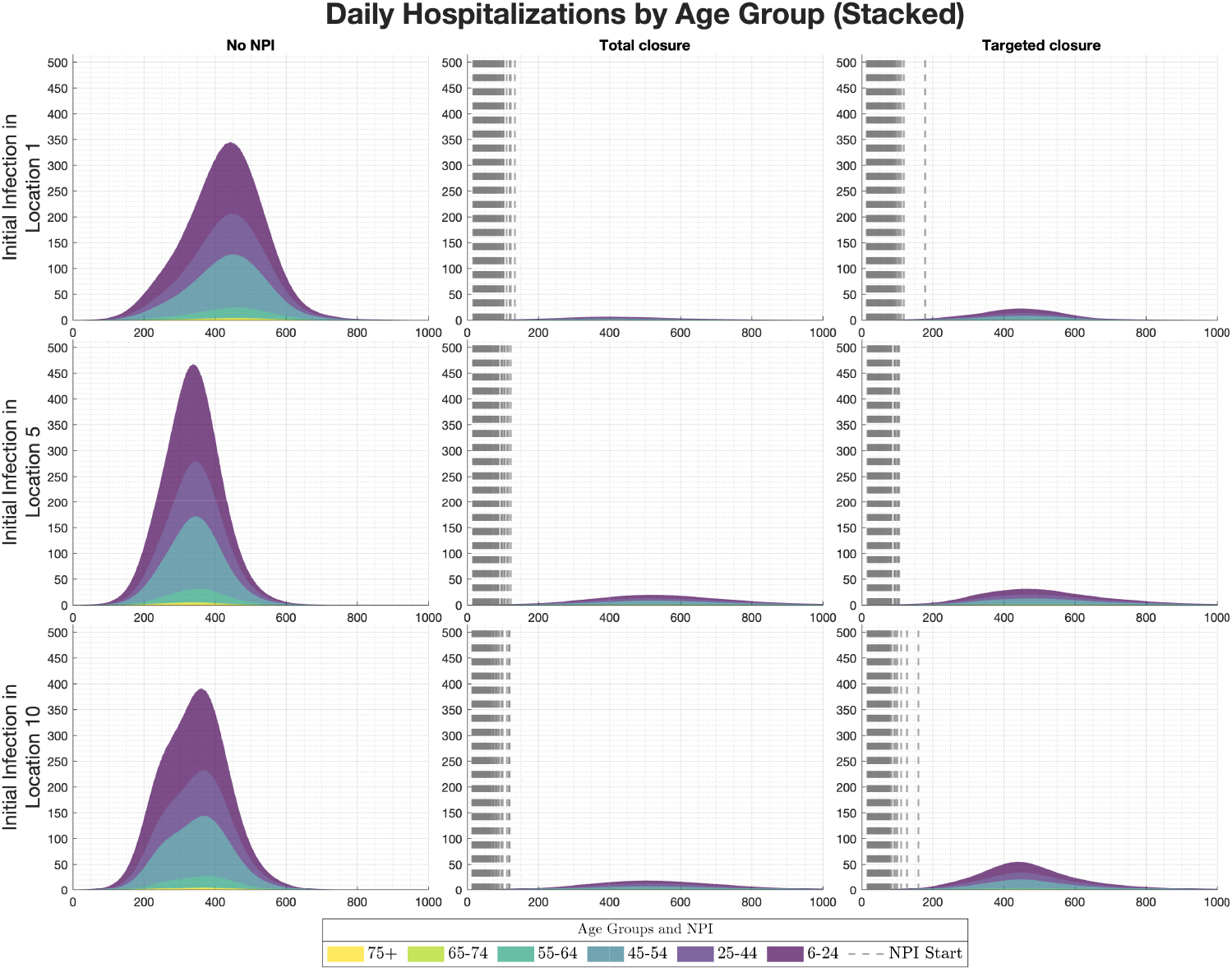
Ebola-like infection: Daily hospitalisations by age group for different scenarios and intervention strategies, shown as stacked area plots.

**Figure 28.**
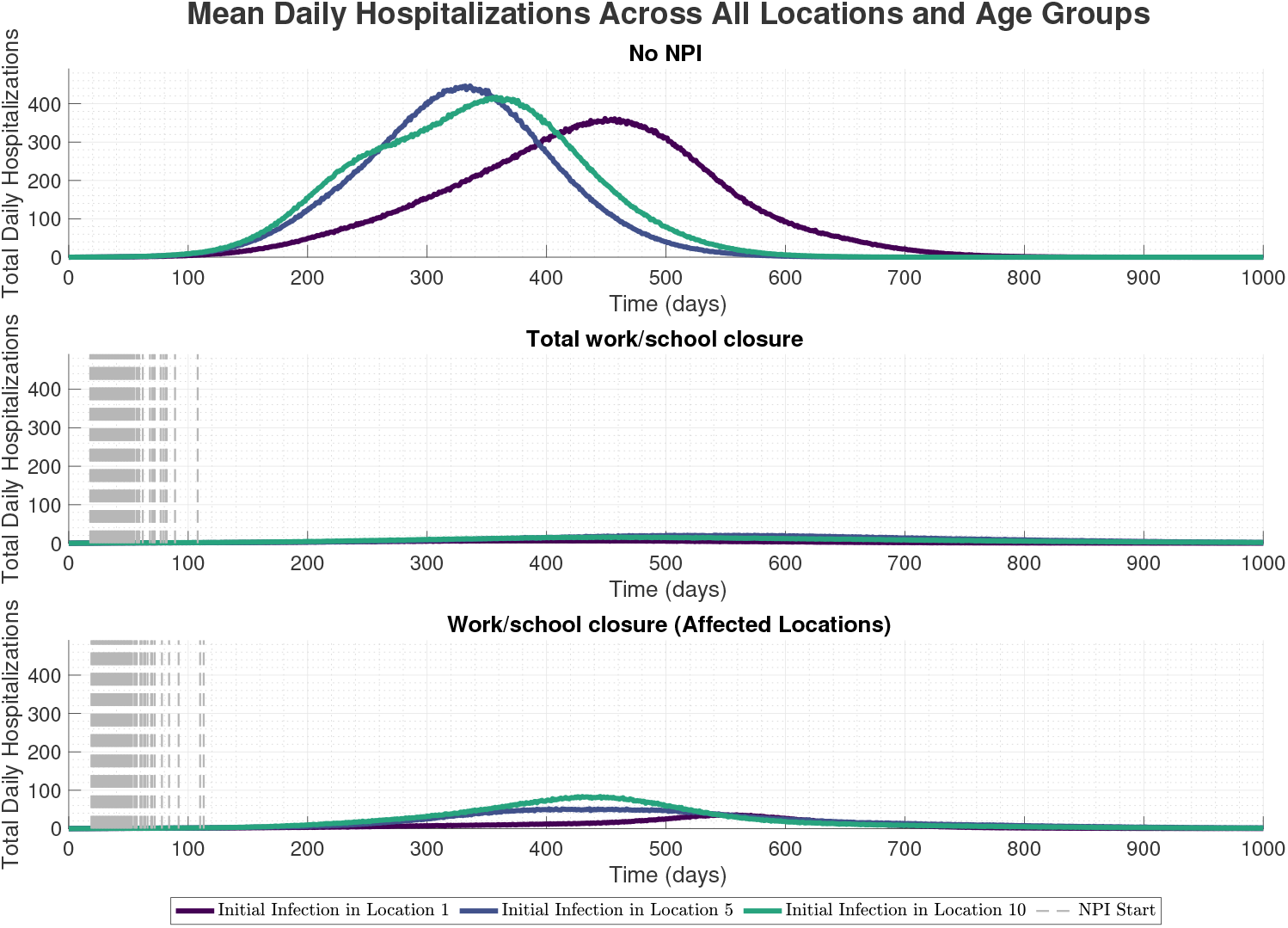
Ebola-like infection: Daily hospitalisations across all locations and age groups for different scenarios and intervention strategies.

**Figure 29.**
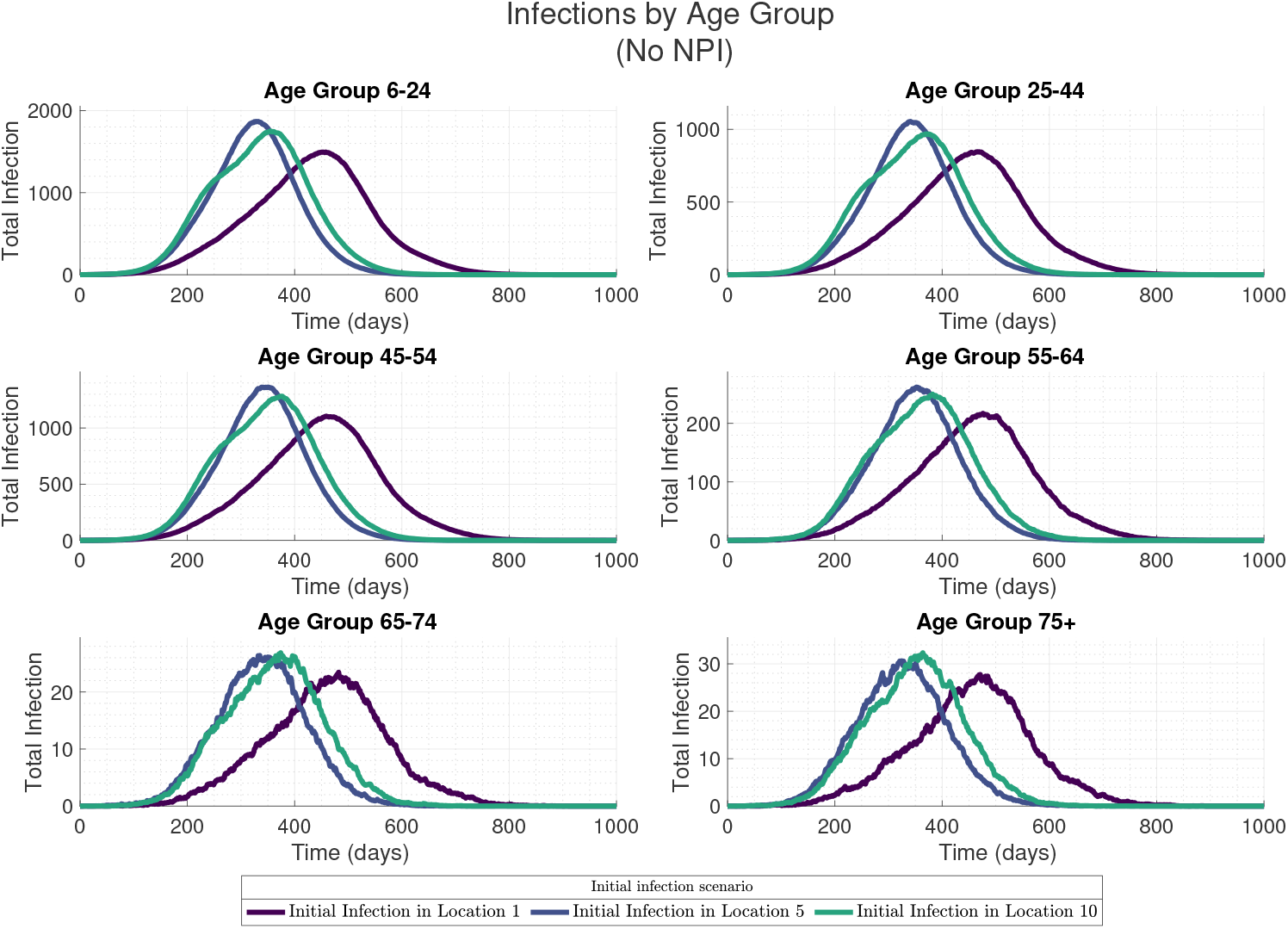
Ebola-like infection: Infections for each age group across all locations with no interventions.

**Figure 30.**
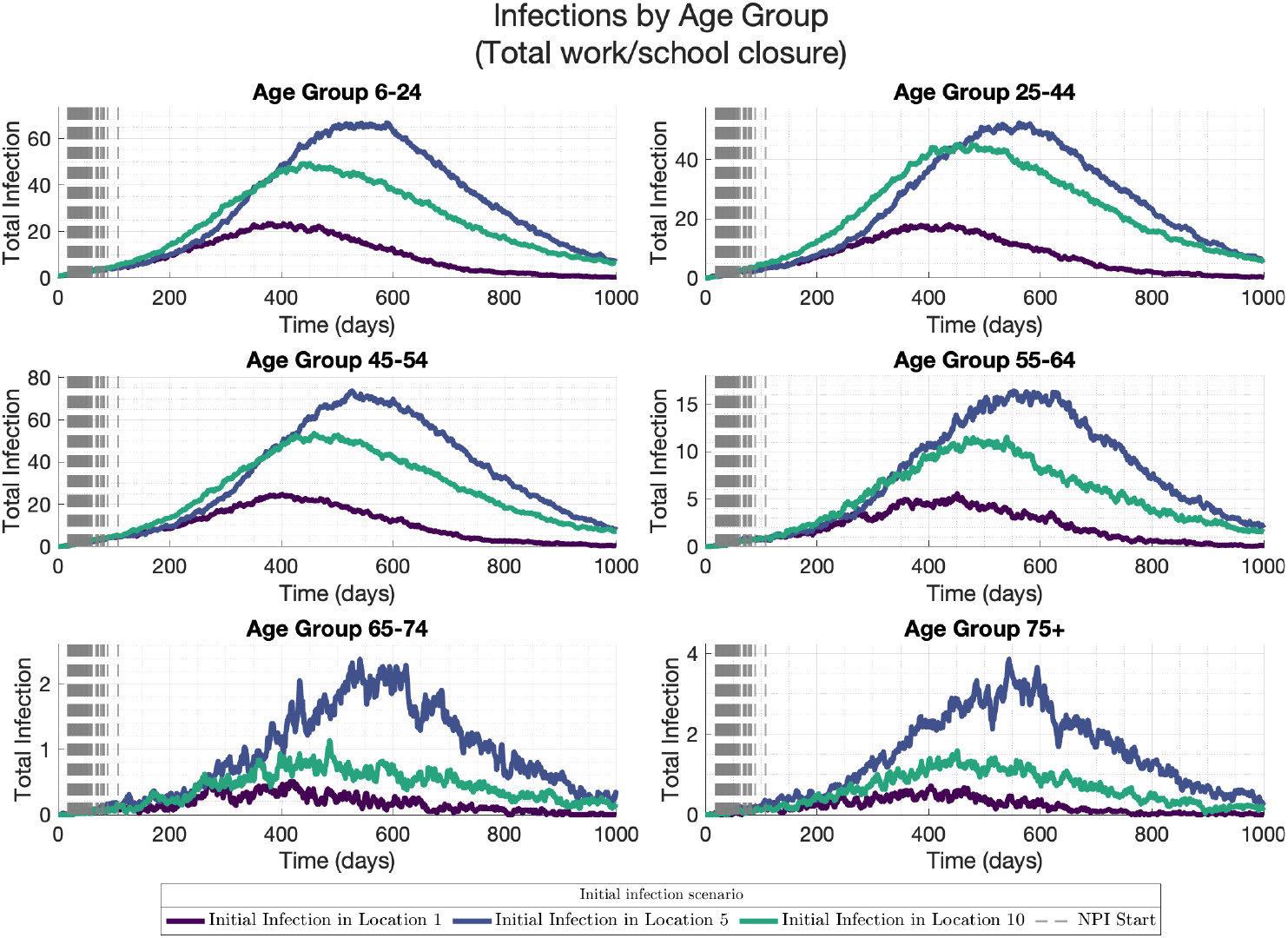
Ebola-like infection: Infections for each age group across all locations with work/school closure in all locations.

**Figure 31.**
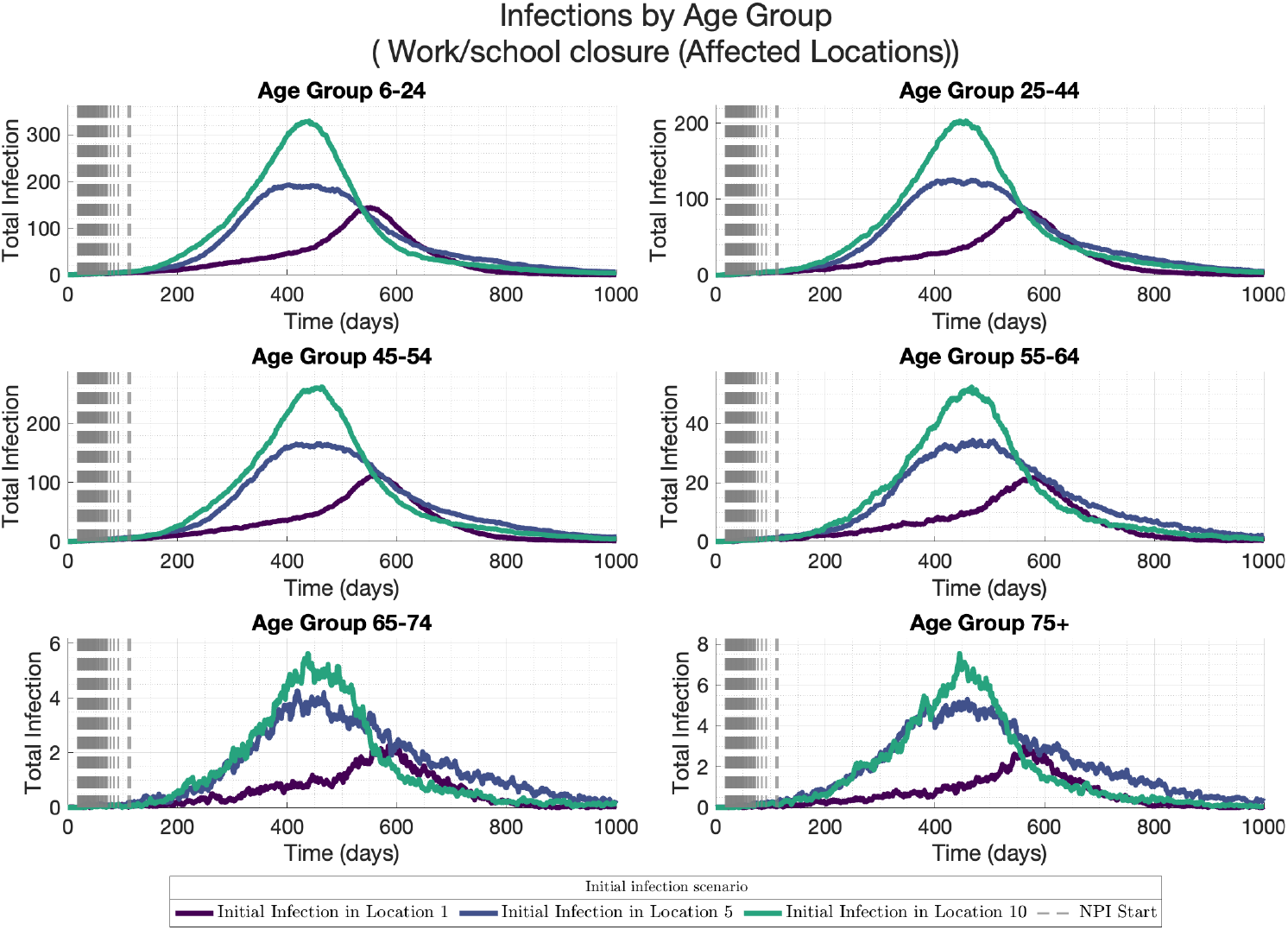
Ebola-like infection: Infections for each age group across all locations with work/school closure in affected locations.

The results of the modelling framework presented in this study for COVID-like and Ebola-like disease transmissions are summarised in Tables 5 and 6. The table presents the probability of large outbreaks and the expected reductions in infections, hospitalisations, and deaths under different intervention strategies and the locations of initial infection. Three seeding locations were considered: Location 10 (large, highly connected), Location 5 (larger, but less connected than Location 10), and Location 1 (smallest among the three example locations, and weakly connected). Intervention strategies included no non-pharmaceutical interventions (No NPI), total work/school closure across all locations (Total), and closure limited to detected locations (Targeted).

**Table 5.** Probability of a large outbreak for COVID-like and Ebola-like transmissions under different seeding locations and intervention strategies (%).

| Seeding location | COVID-like |  |  | Ebola-like |  |  |
| --- | --- | --- | --- | --- | --- | --- |
|  | No NPI | Total | Targeted | No NPI | Total | Targeted |
| Location 10 | 87.0 | 53.3 | 66.0 | 51.2 | 2.8 | 8.2 |
| Location 5 | 84.9 | 17.8 | 37.5 | 48.9 | 0.3 | 3.0 |
| Location 1 | 83.0 | 5.3 | 21.5 | 46.1 | 0.2 | 2.4 |

Notes: “Total” = total work/school closure (all locations); “Targeted” = closure in detected locations only. “Location 10” is large, highly connected, “Location 5” is large, weakly connected, and “Location 1” is small, and weakly connected.

**Table 6.**
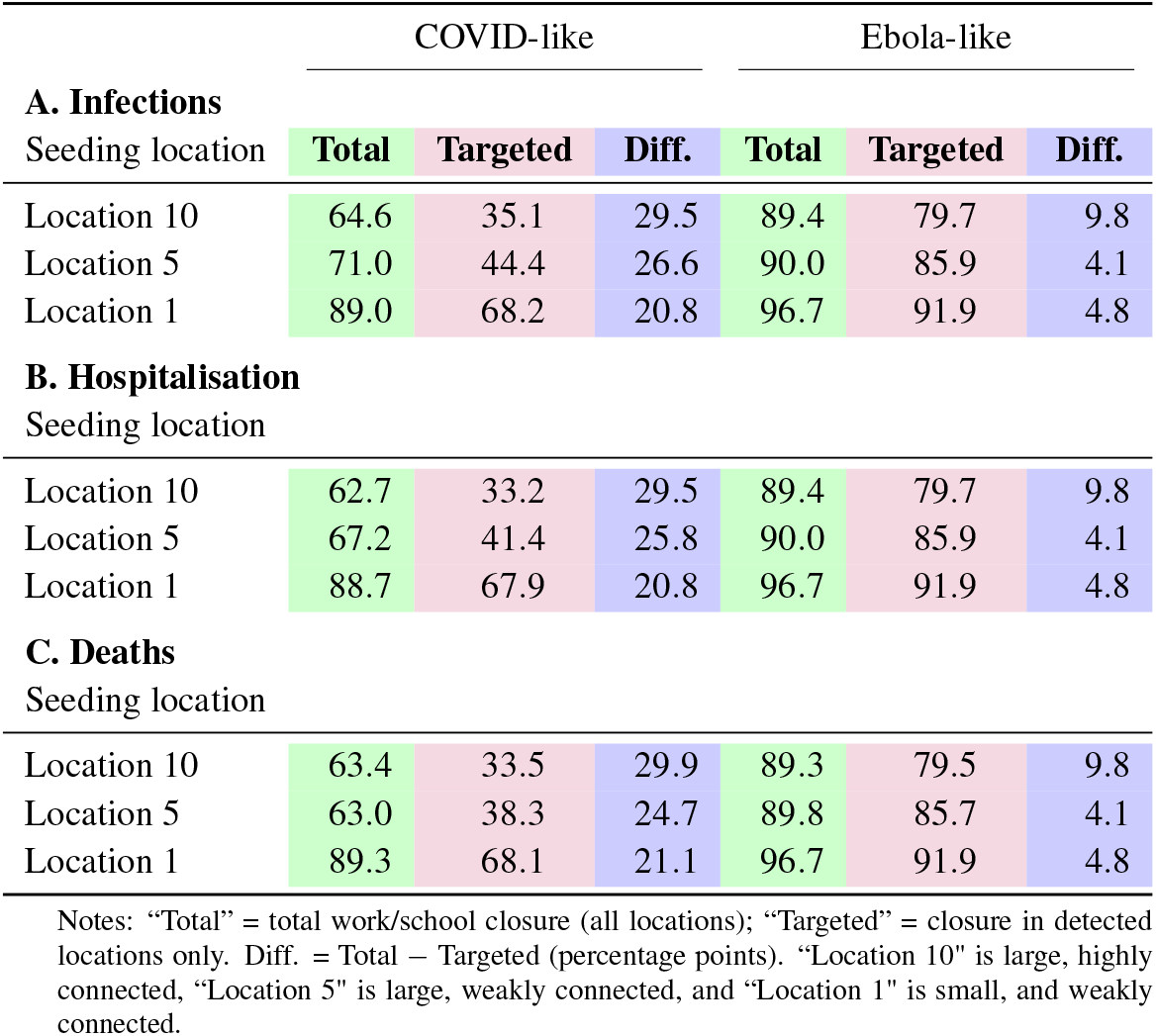
Expected reduction in total infections and deaths under different seeding locations and intervention strategies relative to no-NPI baseline (%).

#### 3.3.1 Probability of Large Outbreaks

Table 5 shows the probability of a large outbreak, defined as an epidemic exceeding a critical threshold of cases and locations (see Methods), for COVID-like and Ebola-like transmissions. If there is no NPI, the chances of a large outbreak of COVID-like transmission are 87.0% if the first infection happened at Location 10, 84.9% if it happened at Location 5, and 83.0% if it happened at Location 1. This shows that there is a high chance of a big outbreak for infection beginning at any of the seeding locations, which is probably because COVID spreads so easily. In contrast, the chances of Ebola-like transmission are lower without any NPIs: 51.2% for Location 10, 48.9% for Location 5, and 46.1% for Location 1. Ebola transmission requires direct contact with the blood, bodily fluid, or skin of an infected person or someone who dies as a result of the disease [46], whereas COVID-19 transmission can occur through air droplets from an infected person. These may likely impact the likelihood of these diseases resulting in major outbreaks.

Implementing interventions lowers the chances of an outbreak happening. The total closure intervention lowers the chance of Covid-like transmission to 53.3% (for an outbreak starting at Location 10), 17.8% (for Location 5), and 5.3% (for Location 1). The biggest drop is seen in places that are less connected. The targeted closure intervention (closing only the places where it was found) is not as effective as the “Total” intervention, with chances of 66.0% (for starting at Location 10), 37.5% (Location 5), and 21.5% (Location 1). For Ebola-like transmission, both interventions are highly effective: the total closure intervention reduces probabilities to 2.8% (Location 10), 0.3% (Location 5), and 0.2% (Location 1), while the targeted closure intervention results in 8.2%, 3.0%, and 2.4%, respectively. These findings highlight that Ebola-like outbreaks are more controllable than COVID-like outbreaks, particularly in less connected locations.

#### 3.3.2 Reduction in Infections, Hospitalisations, and Deaths

Table 6 quantifies the expected reduction in infections, hospitalisations, and deaths relative to the *No NPI* baseline, with differences between *Total* and *Targeted* interventions. For COVID-like transmission, the *Total* intervention shows a reduction in infections by 64.6% (if started at Location 10), 71.0% (for Location 5), and 89.0% (Location 1). The *Targeted* intervention is less effective, with reductions of 35.1%, 44.4%, and 68.2%, respectively. The difference between *Total* and *Targeted* interventions ranges from 20.8% to 29.5%, indicating that total closure consistently outperforms targeted closure, essentially reducing spread particularly when initial infection starts in highly connected locations.

For Ebola-like transmission, both interventions are highly effective. The *Total* intervention reduces infections by 89.4% (Location 10), 90.0% (Location 5), and 96.7% (Location 1), while the *Targeted* intervention achieves 79.7%, 85.9%, and 91.9%, respectively. The differences between interventions are smaller (4.1% to 9.8%), suggesting that targeted interventions are nearly as effective as total closure for Ebola-like diseases, especially if infection starts from the less connected locations.

Similar patterns are observed for hospitalisations and deaths. For COVID-like transmission, reductions in hospitalisations and deaths range from 62.7% to 89.3% for *Total* and 33.2% to 68.1% for *Targeted*, with differences of 20.8% to 29.9%. For Ebola-like transmission, reductions are higher, ranging from 89.3% to 96.7% for *Total* and 79.5% to 91.9% for *Targeted*, with differences of 4.1% to 9.8%. These results underscore the greater efficacy of interventions for Ebola-like diseases, particularly for infection beginning at small, weakly connected locations, where reductions approach 97%.

## 4 Discussion

In this study, we developed and applied a spatial metapopulation epidemic model with recurrent group-switch interactions to investigate the spread and control of epidemics in a heterogeneous population. By integrating the transmission within and outside the household, age, and location-level heterogeneity into a stochastic SEIR framework, the study provides insights into the role of location of initial infection, NPIs, and pathogen-specific characteristics in shaping outbreak trajectories.

The overall epidemic trajectory was largely independent of infection seeding location in the absence of NPIs. Initial infections in highly connected locations (e.g., Location 10) led to higher epidemic peaks and faster spread compared to less connected, smaller populations (e.g., Location 1). This reflects the importance of mobility and commuting patterns in the epidemic trajectory. From a public health perspective, this suggests that surveillance and rapid intervention in highly connected locations could reduce the risk of epidemics, since outbreaks seeded in these areas are more likely to develop.

NPIs can substantially reduce the epidemic size, but this will depend on the location of initial infection, the type of and timing of NPIs. The total closure of schools and workplaces consistently produced the largest reductions in infections, hospitalisations, and deaths, and in stochastic simulations often led to epidemic fade-out. In contrast, the targeted closure in affected locations provided mixed effects depending on the pathogen that was being simulated, indicating that widespread NPIs may be required for some pathogens but may not be required for others if started early in the outbreak, slowing but not preventing multi-location spread, particularly when unreported latent infections or asymptomatic cases were present in Locations without reported cases, at the start of the announcements about interventions. These findings are in agreement with the real-world observations during COVID-19, where interventions implemented with delay allowed widespread community transmission [47]. The results also highlight the trade-off between broad interventions, which are epidemiologically more effective but socially and economically costly, and targeted interventions, which may be more feasible but could be insufficient to fully contain transmission. Policymakers thus face the challenge of balancing immediate health protection with long-term socioeconomic sustainability.

For COVID-19-like infections, analysis by age group revealed that younger adults (6–44) sustained most transmission due to their higher contact rates, whereas older adults experienced fewer infections but higher risks of severe outcomes, including hospitalisation and death. Importantly, interventions indirectly protected older groups by reducing transmission in younger people.

Comparisons of COVID-19–like and Ebola-like infections demonstrated that pathogen characteristics critically shape both epidemic dynamics and intervention effectiveness. COVID-19–like epidemics spread rapidly, reached high peaks, and appeared more difficult to contain than Ebola-like infections, even under total work/school closure. In contrast, Ebola-like infection spread more slowly, allowing interventions such as school and workplace closures to nearly eliminate outbreaks in many simulations. This may be the reason why, although Ebola epidemics have occurred in some parts of West Africa, they have been contained before becoming a global pandemic. These contrasting findings confirm that interventions cannot be applied as one-size-fits-all. For policy, this means that preparedness frameworks must consider pathogen-specific characteristics. While quick and geographically broad NPIs (e.g., total workplace and school closure) may be essential for fast-spreading respiratory pathogens (e.g., COVID-19), more geographically restricted interventions may be sufficient for slower-spreading, usually symptomatic pathogens such as Ebola.

Specifically, the findings in this study provide several key implications: Incorporating stochastic modelling into decision-making: Stochastic simulations capture uncertainty, fade-out probabilities, and variability in epidemic outcomes that deterministic approaches may not reveal. Connectedness of the location of initial case to other locations play important role in the speed of infection and the likelihood of a larger outbreak or pandemic, as initial infection in highly connected locations are more likely to generate large epidemics (or pandemics) and are more difficult to contain. These factors should be accounted for to inform policy, especially, surveillance and public interventions may therefore be best prioritised for transport and commercial hubs. NPIs should be tailored to the speed and transmissibility of the pathogen, informed by models incorporating the factors discussed here. COVID-19–like pathogens require rapid, large-scale interventions, whereas slower pathogens may be controllable with more targeted measures. While total closures are effective in reducing the spread, policymakers must weigh their disruptive impacts, and may need to combine moderate NPIs with case isolation, or vaccination, when available.

Like most epidemiological models, the framework presented in this study made some simplifying assumptions. It assumed that the same age group possesses the same social characteristics (for example, the same cluster sizes and external connections) across all locations, whereas the same age group in different locations may possess varied contact structure. The pathogens simulated here are illustrative examples of an Ebola-like and COVID-like pathogens; they were not fitted to any data, but with hope they reflected the characteristics of potential outbreaks as per the WHO pandemic risk priority diseases list. The number of hospitalizations, deaths, symptomatic and asymptomatic infections have been post-processed from the output of the SEIR model. Detailed dynamics involving these compartments were omitted for simplicity. Moreover, the effects of the transmission of Ebola from the death compartments have not been incorporated. Finally, only two intervention strategies have been simulated in this study, whereas the model structure permits simulation of other NPIs and strategies, for example reducing work/school cluster sizes, reducing external interactions through visit restriction or the approach like bubbles. Although the model framework in this study is capable of implementing all these limitations, these simplifications were made to reduce the volume of work and for faster simulation of the stochastic realisations. These simplifying assumptions could have impacted the quantitative results presented here.

## 5 Conclusion

This study demonstrates that epidemic outcomes in structured metapopulations are shaped by the interplay of stochastic effects, initial location of the infection, intervention strategies, and pathogen characteristics. Deterministic models alone may overestimate outbreaks and potential for pathogen spread, while stochastic approaches highlight the possibility of containment. Modelling to inform public health policy should consider the choice of stochastic modelling framework, since it is more realistic, and also the consider integrating more realistic factors of mobility and interaction (e.g., non-random mixing, multi-phase interaction and utilising the origin-destination data to estimate probability of inter-location epidemic spread) into epidemic models. Public health responses should therefore be adaptive: prioritising rapid, broad interventions for fast-spreading pathogens, tailoring strategies to pathogen-specific characteristics, and focusing early containment efforts in central hubs. The model presented here is flexible and could be adapted to different pathogens and other NPIs targeting different locations or sociodemographic groups. These insights contribute to a more nuanced understanding of how spatial heterogeneity and recurrent group-switch interactions influence epidemic spread and context-specific policies to mitigate future epidemics and pandemics.

## Data Availability

All data produced in the present study are available upon reasonable request to the corresponding authour

## Acknowledgements

MLS was fully funded by the Institute for Global Pandemic Planning, Warwick Medical School, University of Warwick UK.

## Author contributions statement

Conceptualization: M. L. S.

Methodology: M. L. S.

Supervision: A.S. & K. S. R.

Software: K. S. R. & M. L. S.

Visualization: K S. R & M. L. S.

Writing – original draft: M. L. S.

Writing – review & editing: A. S., K. S. R. & M. L. S.

## Data availability

MATLAB code shall be accessible via GitHub upon publication of the manuscript.

## Ethics declarations

This study focuses on the theoretical development of a mathematical model for the spread of a hypothetical infectious disease. No data was collected and analysed.

## Additional information

### Competing interests

The authors declare no competing interests. For open access, the authors have applied a Creative Commons Attribution (CC-BY) licence to any Author Accepted Manuscript version arising from this submission.

## Notes

### Competing Interest Statement

The authors have declared no competing interest.

### Author Declarations

This study focuses on the theoretical development of a mathematical model for the spread of a hypothetical infectious disease.

## References

1. WHO. Prioritizing diseases for research and development in emergency contexts. https://www.who.int/activities/prioritizing-diseases-for-research-and-development-in-emergency-contexts (2021). [Accessed 31-January-2023].

2. Nyakarahuka, L. et al. Ten outbreaks of Rift Valley fever in uganda 2016-2018: epidemiological and laboratory findings. Int. J. Infect. Dis. 79, 4 (2019).

3. El Mamy, A. B. O. et al. Unexpected Rift Valley fever outbreak, Northern Mauritania. Emerg. Infect. Dis. 17, 1894 (2011).

4. Buseh, A. G., Stevens, P. E., Bromberg, M. & Kelber, S. T. The Ebola epidemic in west africa: Challenges, opportunities, and policy priority areas. Nurs. Outlook 63, 30–40 (2015).

5. Spiteri, G. et al. First cases of coronavirus disease 2019 (COVID-19) in the WHO European Region, 24 january to 21 february 2020. Eurosurveillance 25, 2000178 (2020).

6. Paul, E., Brown, G. W., Bell, D., von Agris, J. M. & Ridde, V. Royal society report: what would a comprehensive evaluation suggest about non-pharmaceutical interventions during COVID-19? Critical Public Heal. 34, 1–10 (2024).

7. Maffioli, E. M. How is the world responding to the novel coronavirus disease (COVID-19) compared with the 2014 West African Ebola epidemic? the importance of China as a player in the global economy. The Am. J. Trop. Medicine Hyg. 102, 924 (2020).

8. Chang, C. et al. The novel h1n1 influenza a global airline transmission and early warning without travel containments. Chin. Sci. Bull. 55, 3030–3036 (2010).

9. Neal, P. A household SIR epidemic model incorporating time of day effects. J. Appl. Probab. 53, 489–501 (2016).

10. Hilton, J. & Keeling, M. J. Incorporating household structure and demography into models of endemic disease. J. Royal Soc. Interface 16, 20190317 (2019).

11. Hilton, J. et al. A computational framework for modelling infectious disease policy based on age and household structure with applications to the COVID-19 pandemic. PLoS Comput. Biol. 18, e1010390 (2022).

12. Garnier, J. & Lafontaine, P. Dispersal and good habitat quality promote neutral genetic diversity in metapopulations. Bull. Math. Biol. 83, 1–51 (2021).

13. Rao, S., Muyinda, N. & De Baets, B. Stability analysis of the coexistence equilibrium of a balanced metapopulation model. Sci. Reports 11, 1–15 (2021).

14. Kelly Jr, M. R., Tien, J. H., Eisenberg, M. C. & Lenhart, S. The impact of spatial arrangements on epidemic disease dynamics and intervention strategies. J. biological dynamics 10, 222–249 (2016).

15. Nagatani, T., Ichinose, G. & Tainaka, K.-i. Epidemics of random walkers in metapopulation model for complete, cycle, and star graphs. J. Theor. Biol. 450, 66–75 (2018).

16. Silal, S. P., Little, F., Barnes, K. I. & White, L. J. Predicting the impact of border control on malaria transmission: a simulated focal screen and treat campaign. Malar. journal 14, 1–14 (2015).

17. Anderson, S. et al. An agent-based metapopulation model simulating virus-based biocontrol of Heterodera glycines. J. Nematol. 50, 79 (2018).

18. Balcan, D. et al. Modeling the spatial spread of infectious diseases: The global epidemic and mobility computational model. J. computational science 1, 132–145 (2010).

19. Hinch, R. et al. Estimating SARS-CoV-2 variant fitness and the impact of interventions in england using statistical and geo-spatial agent-based models. Philos. Transactions Royal Soc. A 380, 20210304 (2022).

20. Winkelmann, S., Zonker, J., Schütte, C. & Conrad, N. D. Mathematical modeling of spatio-temporal population dynamics and application to epidemic spreading. Math. Biosci. 336, 108619 (2021).

21. Cota, W., Soriano-Paños, D., Arenas, A., Ferreira, S. C. & Gómez-Gardeñes, J. Infectious disease dynamics in metapopulations with heterogeneous transmission and recurrent mobility. New J. Phys. 23, 073019 (2021).

22. Soriano-Paños, D., Ghoshal, G., Arenas, A. & Gómez-Gardeñes, J. Impact of temporal scales and recurrent mobility patterns on the unfolding of epidemics. J. Stat. Mech. Theory Exp. 2020, 024006 (2020).

23. Gómez, S., Arenas, A., Borge-Holthoefer, J., Meloni, S. & Moreno, Y. Discrete-time Markov chain approach to contact-based disease spreading in complex networks. EPL (Europhysics Lett. 89, 38009 (2010).

24. Gómez-Gardeñes, J., Soriano-Panos, D. & Arenas, A. Critical regimes driven by recurrent mobility patterns of reaction– diffusion processes in networks. Nat. Phys. 14, 391–395 (2018).

25. Soriano-Paños, D., Lotero, L., Arenas, A. & Gómez-Gardeñes, J. Spreading processes in multiplex metapopulations containing different mobility networks. Phys. Rev. X 8, 031039, DOI: 10.1103/PhysRevX.8.031039 (2018).

26. Huang, J. & Chen, C. Metapopulation epidemic models with a universal mobility pattern on interconnected networks. Phys. A: Stat. Mech. its Appl. 591, 126692 (2022).

27. Valgañón, P., Soriano-Paños, D., Arenas, A. & Gómez-Gardeñes, J. Contagion–diffusion processes with recurrent mobility patterns of distinguishable agents. Chaos: An Interdiscip. J. Nonlinear Sci. 32, 043102 (2022).

28. Reyna-Lara, A., Soriano-Paños, D., Arenas, A. & Gómez-Gardeñes, J. The interconnection between independent reactive control policies drives the stringency of local containment. Chaos, Solitons & Fractals 158, 112012 (2022).

29. Soriano-Paños, D. et al. Modeling communicable diseases, human mobility, and epidemics: A review. Annalen der Physik 534, 2100482 (2022).

30. Gómez, S., Fernández, A., Meloni, S. & Arenas, A. Impact of origin-destination information in epidemic spreading. Sci. reports 9, 2315 (2019).

31. Smah, M. L., Seale, A. C. & Rock, K. S. Recurrent group-switch interactions in heterogeneous population epidemic modelling. medRxiv 2025–11 (2025).

32. Coletti, P. et al. A data-driven metapopulation model for the Belgian COVID-19 epidemic: assessing the impact of lockdown and exit strategies. BMC infectious diseases 21, 503 (2021).

33. Chinazzi, M. et al. A multiscale modeling framework for Scenario Modeling: Characterizing the heterogeneity of the COVID-19 epidemic in the US. Epidemics 47, 100757 (2024).

34. Su, Q. et al. Assessing the burden of congenital rubella syndrome in China and evaluating mitigation strategies: a metapopulation modelling study. The Lancet Infect. Dis. 21, 1004–1013 (2021).

35. Metcalf, C. J. E., Munayco, C. V., Chowell, G., Grenfell, B. T. & Bjørnstad, O. N. Rubella metapopulation dynamics and importance of spatial coupling to the risk of congenital rubella syndrome in Peru. J. Royal Soc. Interface 8, 369–376 (2011).

36. Smah, M. L., Seale, A. C. & Rock, K. S. Modelling semi-random human mixing: A modified force of infection in population-level epidemic modelling. medRxiv 2025–08 (2025).

37. Smah, M. L., Seale, A. C. & Rock, K. S. Semi-random mixing epidemic model: Integrating explicit household and non-household interactions. medRxiv 2025–11 (2025).

38. Mossong, J. et al. POLYMOD social contact data. http://www.socialcontactdata.org (2017). Accessed on September 9, 2023.

39. Nomis - Official Labour Market Statistics. https://www.nomisweb.co.uk/query/select/getdatasetbytheme.asp?opt=3&theme=&subgrp=. Accessed on August 1, 2023.

40. Ward, T. et al. The real-time infection hospitalisation and fatality risk across the COVID-19 pandemic in england. Nat. Commun. 15, 4633 (2024).

41. Wang, B. et al. Asymptomatic SARS-CoV-2 infection by age: a global systematic review and meta-analysis. The Pediatr. Infect. Dis. J. 42, 232–239 (2023).

42. European Centre for Disease Prevention and Control. Factsheet about Ebola disease. https://www.ecdc.europa.eu/en/infectious-disease-topics/ebola-virus-disease/facts/factsheet-about-ebola-disease (2023). Accessed: 2025-08-22.

43. Funk, S. et al. The impact of control strategies and behavioural changes on the elimination of Ebola from lofa county, Liberia. Philos. Transactions Royal Soc. B: Biol. Sci. 372, 20160302 (2017).

44. Team, W. E. R. Ebola virus disease among children in west africa. New Engl. J. Medicine 372, 1274–1277 (2015).

45. Glynn, J. R. et al. Asymptomatic infection and unrecognised Ebola virus disease in Ebola-affected households in Sierra Leone: a cross-sectional study using a new non-invasive assay for antibodies to Ebola virus. The Lancet infectious diseases 17, 645–653 (2017).

46. Rewar, S. & Mirdha, D. Transmission of Ebola virus disease: an overview. Annals global health 80, 444–451 (2014).

47. Pei, S., Kandula, S. & Shaman, J. Differential effects of intervention timing on COVID-19 spread in the United States. Sci. advances 6, eabd6370 (2020).

